# Statins and antidepressants induce similar *in vitro* gene expression responses

**DOI:** 10.1101/2022.03.27.22273017

**Authors:** Jiayue-Clara Jiang, Chenwen Hu, Andrew M McIntosh, Sonia Shah

## Abstract

**Background:** Based on observational studies and small-scale randomized controlled trials, it is uncertain whether cholesterol-lowering statins have any beneficial or adverse effects on depressive symptoms. In this study we investigate this question using a genomics approach.

**Methods:** To compare the pharmacological effects of statin and antidepressant exposure and identify commonly perturbed biological pathways, we interrogated Connectivity Map (CMap), a database of gene expression signatures from drug-treated human cell lines. We used Mendelian randomization, a statistical genomics approach, to investigate the potential causal on-target (HMGCR inhibition) and off-target effects of statin exposure on depression, depressive symptoms, and traits related to the shared pathways identified from CMap analysis.

**Results:** Compounds inducing highly similar gene expression responses to statins (as indicated by an average CMap connectivity score with statins > 90) were enriched for antidepressants (12 out of 38 antidepressants; p < 1E-05). Genes perturbed in the same direction by both statins and antidepressants were significantly enriched for diverse cellular and metabolic pathways, and various immune activation, development and response processes. Genetically proxied HMGCR inhibition was significantly associated with monocyte and platelet-related metrics.

**Conclusions:** Our study is the first to directly compare gene expression responses to statins and antidepressants, demonstrating perturbation of shared immune pathways. We further show that statin exposure is strongly associated with alterations in monocyte and platelet measures, both of which have previously been implicated in depression. Our findings warrant further investigation into the use of statins for treating depression, particularly in patients with raised blood biomarkers of inflammation.

## Introduction

Depression is one of the most common mental disorders and a leading cause of non-fatal health loss. Pharmacological treatments for depression involve the use of antidepressants to alleviate symptoms; however, conventional antidepressants often show latency in treatment response and limited remission rates (1). Furthermore, there is poor mechanistic understanding of antidepressant actions, making it difficult to develop treatments for depression.

Drug repurposing is the process of identifying novel therapeutic effects of existing drugs, and represents an efficient and cost-effective approach for developing new therapeutic treatments (2). Genome-wide association studies (GWAS) have guided the drug repurposing strategies for several diseases, with a notable example being the repurposing of a psoriasis treatment, Ustekinumab, for Crohn’s disease (2, 3). Another computational approach for evaluating drug repurposing potential is signature matching, where the molecular changes (e.g. gene or protein expression) in response to drug candidate exposure are compared against drugs approved for treating the disease. The underlying assumption is that if two drugs have shared pharmacological effects, they likely induce highly similar molecular responses (2).

3-Hydroxy-3-Methylglutaryl Coenzyme A Reductase (HMGCR) inhibitors, or collectively known as statins, effectively lower low-density lipoprotein cholesterol (LDL-C) levels, and are widely prescribed for treating cardiovascular diseases (4, 5). There is conflicting evidence on the anti-depressive effect of statins from observational studies and randomized controlled trials (RCT) (6). This inconsistency in findings may be due to differences in the chemical and pharmacokinetic properties of statin compounds studied (7), small sample sizes, varying follow-up times, and unmeasured confounders. Several mechanisms have been proposed to explain the anti-depressive effects of statins, including those mediated through LDL-C lowering and reduction of atherosclerosis, downregulation of immune activation and pro-inflammatory cytokines such as C-reactive protein (CRP) (both dependent and independent of their lipid-lowering activity), and reduction in glucocorticoid concentrations (6). However, the exact relationship between statins and depression remains largely unknown. In this study, we aimed to investigate the anti-depressive effects of statins by mining the Connectivity Map (CMap) database of gene expression signatures from drug-treated human cell lines. We also performed Mendelian randomization (MR) analysis, using genetic variants associated with drug target gene expression as instruments for drug exposure, to assess the causal association between statin exposure and outcomes of interest, including depression risk and related traits. MR can overcome some of the issues of observational studies, such as unmeasured confounders and sample size issues, since genetic studies nowadays often include hundreds of thousands of individuals.

## Methods and Materials

### CMap gene expression signatures

Each CMap gene expression signature consists of the gene expression changes (z-scores) of 978 directly measured “landmark” genes and 11 350 inferred genes (Supplementary Methods) (8). We retrieved the z-scores of CMap signatures from the GEO repository (GSE92742) (accessed on July 8^th^, 2021). We selected the signatures of statins in the HA1E kidney cell line for analysis, due to the greatest concordance and specificity in gene expression changes amongst different statin compounds (Supplementary Figure 1) (Supplementary Methods). A total of 7 statins (atorvastatin, fluvastatin, lovastatin, mevastatin, pravastatin, rosuvastatin, and simvastatin) were profiled in HA1E cells under the selected perturbational conditions (time = 24 h, dose = 10 *μ*M), with pravastatin discarded from further analysis due to low correlation with other statins (Supplementary Figure 2). We also retrieved the gene expression signatures of antidepressants, and included alvespimycin (heat shock protein inhibitor) and sirolimus (mTOR inhibitor) as control compounds with no known anti-depressive or cholesterol-lowering effects. The CMap signatures analyzed in this study are summarized in Supplementary Table 1.

### Connectivity scores

The CMap platform computes a “connectivity” score (Tau) as a measure of similarity between a query gene expression signature, in this case the signature for statin exposure, and a reference signature (here we are interested in signatures of antidepressants) (Supplementary Methods) (8). The Tau scores range between −100 and 100, and the magnitude of the score corresponds to the magnitude of similarity or dissimilarity in the gene expression signatures. In general, a Tau score greater than 90 is used as a threshold for identifying compounds with potentially shared pharmacological effects (8).

For each statin, we constructed a query signature using 50 most up-regulated and 50 most down-regulated landmark genes defined by z-scores (referred as the top 50 gene set), which is the default criteria used by CMap to generate Tau scores. The query signature for each statin was compared against all gene expression signatures in the CMap database (LINCS version 1.0), and Tau scores were retrieved using the CLUE platform. As a sensitivity analysis, we repeated the connectivity analysis using the top 100 and 150 landmark genes. The similarity between specific compound signatures was further evaluated by investigating the correlation of the z-scores of all 12 328 genes profiled in each CMap signature (Supplementary Methods).

### Pathway enrichment analysis for genes perturbed by statins

We used z-score > 1 and z-score < −1 to identify genes up-regulated and down-regulated, respectively, in response to drug exposure. To identify Gene Ontology (GO) biological processes perturbed by statins, functional enrichment analysis by gProfiler2 (version 0.2.0) (Ensembl 104) was performed for up-regulated and down-regulated genes in R (9, 10), using all 12 328 CMap-profiled genes as the background gene list and a gSCS-corrected p-value < 0.05 to indicate statistical significance. The gSCS multiple testing correction method accounts for the overlapping and hierarchically related nature of GO functional terms and gives a more stringent significance threshold (9, 10). Given the many overlapping GO terms, we categorized the significantly enriched biological process terms into higher-level ancestor terms (Supplementary Methods). As a sensitivity analysis, we repeated pathway enrichment analysis using different z-score thresholds (|z| of 1.5 and 2) to define differentially expressed genes (Supplementary Table 2).

### Enrichment of antidepressants amongst high-connectivity compounds

The gene expression signatures of 38 antidepressants (ATC code: N06A) are profiled in HA1E cells (Supplementary Table 3 and Supplementary Methods). We performed a chi-square test to determine enrichment of antidepressants among compounds with high positive connectivity with statins (average Tau > 90). A chi-square p-value < 0.05 defined statistical significance.

### Pathway enrichment analysis for genes perturbed by both statins and antidepressants

Out of the antidepressants with highly similar gene expression signatures to statins in HA1E cells, we selected the top five antidepressants (paroxetine, sertraline, nortriptyline, trimipramine, and desipramine) that demonstrated the highest average connectivity with the six statins for detailed analysis (Supplementary File 1). We identified genes that were differentially expressed (as above) in response to both statins and antidepressants. We performed pathway enrichment analysis on genes perturbed in the same and opposite direction by each pair of statin and antidepressant (Supplementary Methods).

### Mendelian randomization analysis

MR is a statistical genetics approach used to estimate the causal effect of an exposure on an outcome of interest (detailed description in Supplementary Methods). Statins are potent inhibitors of the HMGCR protein, and statin-mediated HMGCR inhibition is the primary pathway underlying their lipid-lowering effects (4, 5). Selective statins have also been reported to exert *in vitro* off-target inhibition of Integrin Alpha-L (ITGAL) and Histone Deacetylase 2 (HDAC2) (details provided in Supplementary Methods), though this off-target inhibition is not related to the cholesterol-lowering properties of statins. We used MR analysis to investigate both on-target and off-target effects of statins.

### Genetic instruments for statin target inhibition

We used expression quantitative trait loci (eQTL) of *HMGCR, ITGAL* and *HDAC2* expression in blood (eQTLGen (11)) to proxy for statin exposure (Supplementary Methods and Supplementary Table 4). We used eQTLs with F-statistic > 10, indicating strong instruments. To investigate the on-target effect of statins, we selected rs12916, which is a previously validated instrument for HMGCR inhibition, and a strong cis-eQTL for *HMGCR* expression in blood (p = 1.5E-36). Given the relevance of the tissue for depression, we performed additional analysis using a strong eQTL (rs17671591; p = 2.5E-05) for *HMGCR* expression in the brain prefrontal cortex (PsychENCODE (12)), which is in moderate linkage disequilibrium (LD) (r^2^ = 0.6) with rs12916 amongst European individuals. To make inferences about the off-target effects of statins, we used the most significant blood eQTLs as genetic proxies for ITGAL (rs11574938; p = 7.9E-150) and HDAC2 (rs9481408; p = 4.1E-07) inhibition.

### Outcome GWAS data

Publicly available GWAS summary statistics, derived from European cohorts, were obtained for 29 immune and disease traits, including haematological traits, interleukin-6 (IL6) and CRP levels, depression, and depression-related symptoms (neuroticism, worrying and depressed affect). In addition, we included several proof-of-principle traits previously linked to statin use, including blood lipid levels (high-density lipoprotein cholesterol (HDL-C), LDL-C, and triglycerides (TG)), body mass index (BMI), coronary artery disease (CAD), and type II diabetes (T2D). Details of each GWAS are provided in Supplementary Table 5.

### SMR analysis

We used SMR (13) (version 0.710) to evaluate the association of genetically proxied statin on-target and off-target inhibition with various traits. We used blood (*HMGCR, ITGAL* and *HDAC2*) or brain (*HMGCR*) gene expression as exposure, and disease and haematological traits as outcome. GWAS summary-data-based MR methods require a reference dataset for LD estimation, and we used a random sample of 10 000 European individuals from the UK Biobank as the LD reference. A threshold of p < 0.00057 (after multiple testing correction for three statin target genes and 29 traits of interest) defined statistical significance for the SMR test. We performed the heterogeneity in dependent instruments (HEIDI) test as a sensitivity analysis (13), where a HEIDI p-value > 0.01 indicated that the observed exposure-outcome association was mediated through one causal single nucleotide polymorphism (SNP), rather than via LD between separate SNPs (linkage scenario) (Supplementary Methods).

## Results

### Statins induce extensive *in vitro* gene expression changes

We observed a high correlation in gene expression changes across six statins (atorvastatin, fluvastatin, lovastatin, mevastatin, rosuvastatin, and simvastatin) in HA1E cells (Supplementary Figure 3), supported by high connectivity scores (median Tau = 99.95). The high concordance amongst statin-induced signatures was consistently observed regardless of the number of differentially expressed genes used to compute the connectivity profile (Supplementary Figure 4), demonstrating the robustness of these signatures.

### Statins induce gene expression changes in lipid and immune-associated pathways

We performed functional enrichment analysis on genes up-regulated (z-score > 1) and down-regulated (z-score < −1) by statins in HA1E cells (Supplementary Figure 5 and Supplementary File 2). A total of 366 GO biological process terms were significantly enriched amongst the up- and down-regulated genes induced by at least one statin. Cellular processes, including various DNA replication and RNA processing pathways, were the most enriched category of biological processes (Supplementary Figure 5). Statins also induced extensive perturbations in metabolism, biological regulation, localization, response to stimulus and immune processes (Supplementary Figure 5). As expected from their lipid-lowering effects, statins induced widespread expression changes amongst genes annotated with lipid metabolic pathways. We confirmed the reproducibility of pathway enrichment analysis results using more stringent thresholds for defining differential gene expression (|z| > 1.5 or 2) (Supplementary Figure 6). We investigated the gene expression signatures of statins in HepG2 (liver) and MCF7 (breast) cell lines, and found overall positive correlation across different cell lines in the gene expression changes induced by statin exposure, both globally as well as amongst lipid and immune-associated genes (Supplementary Figure 7).

### Immune pathways are perturbed by statins and antidepressants in concordant direction

The CMap connectivity analysis identified 201 out of 2770 (7.3%) non-statin compounds with an average Tau score greater than 90, indicating strong positive correlation to statin-induced gene expression changes (Supplementary File 1). 14 compounds had a Tau score lower than −90, suggesting opposite transcriptomic impacts (Supplementary File 3). Antidepressants were significantly enriched amongst compounds with highly similar gene expression signatures to statins (p < 1E-05), with 12 out of the 38 (31.6%) antidepressants profiled by the CMap database having an average Tau score greater than 90 with statins (Figure 1A and Supplementary File 4). We performed functional enrichment analysis on three selective monoamine reuptake inhibitors (desipramine, trimipramine and nortriptyline) (ATC code: N06AA) and two selective serotonin reuptake inhibitors (paroxetine and sertraline) (ATC code: N06AB) that showed the highest average connectivity to the six statins in HA1E cells (Figure 1A and Supplementary Figure 8).

**Figure 1.**
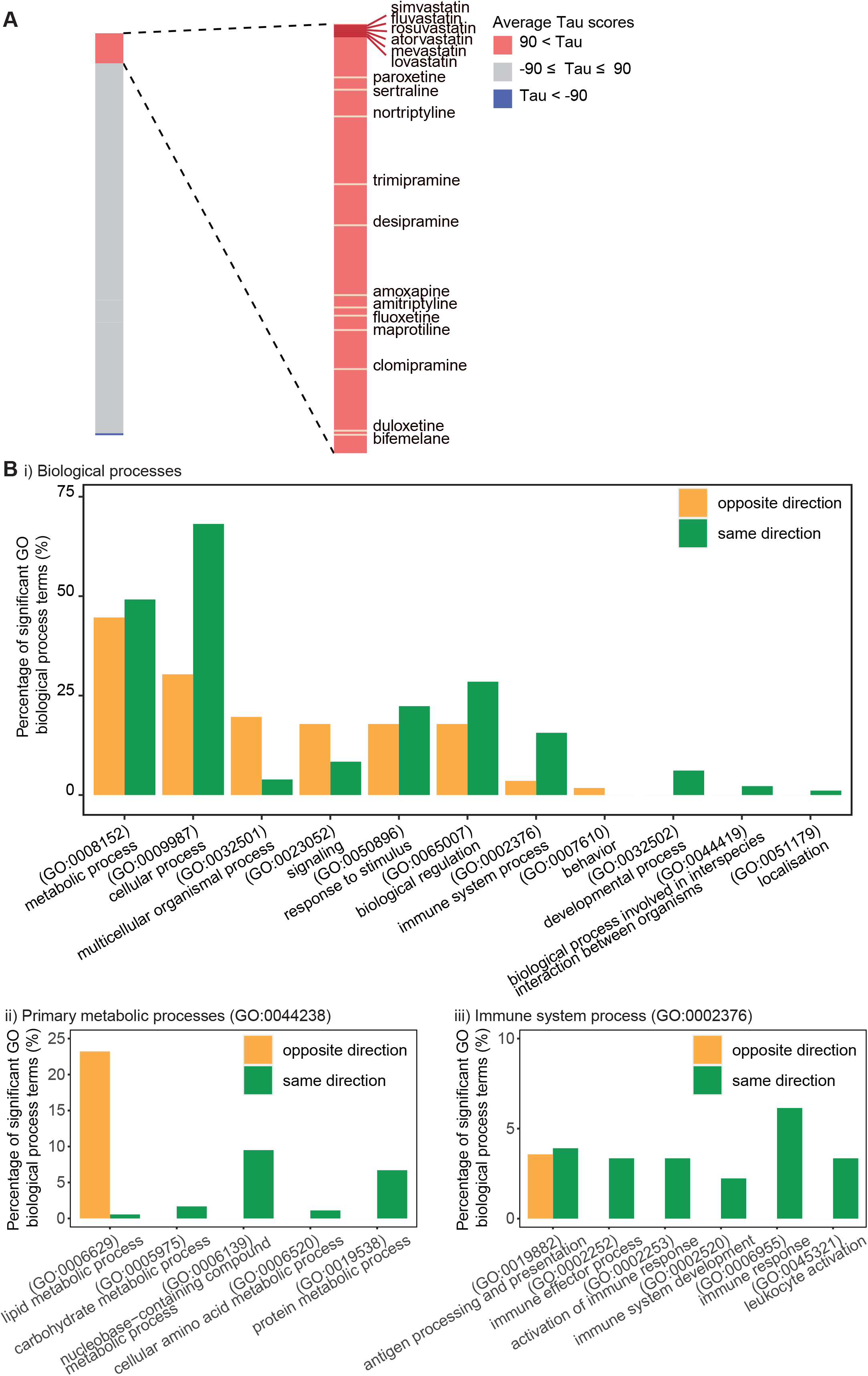
Statins and antidepressants induced similar transcriptomic responses in HA1E cells. (**A**) Statins and antidepressants ranked by connectivity (Tau) scores (averaged across six statins). Lengths of the colored bars represent the number of compounds with the corresponding average Tau scores (red: Tau > 90; grey: −90 ≤ Tau ≤ 90; blue: Tau < −90). The ranking of antidepressants with an average Tau score higher than 90 are shown. (**B**) Biological processes identified as significantly enriched amongst genes perturbed by both statins and antidepressants. A total of 179 and 56 Gene Ontology (GO) biological process terms were identified to be significantly enriched amongst genes perturbed in the same (green) and opposite (yellow) directions respectively, and categorized into high-level ancestor terms. Bar graph shows the percentages of significant GO biological process terms annotated with (i) high-level biological process ancestor terms, (ii) child terms of primary metabolic process (GO:0044238) (a child term of “metabolic process”), and (iii) child terms of immune system process (GO:0002376).

We identified 179 and 56 biological processes to be significantly enriched in genes perturbed in the same and opposite directions (Supplementary Table 6), respectively, by at least one statin-antidepressant pair (Figure 1B, Supplementary File 5 and Supplementary File 6). Notably, lipid metabolic processes were mostly enriched amongst genes perturbed in the opposite direction, supporting previous findings on the cholesterol-increasing effects of antidepressants (Supplementary Figure 9) (14). In comparison, genes perturbed in the same direction were, to a higher extent, functionally involved in cellular processes and biological regulation processes, such as cell cycle regulation, RNA processing, and cellular component biogenesis. Furthermore, genes perturbed in the same direction were enriched in diverse immune system processes involved in both innate and adaptive immunity, such as granulocyte activation, myeloid leukocyte activation, T cell receptor signaling, as well as antigen processing and presentation (Supplementary File 5). Pathways involving inflammatory cytokines, such as interleukin-1 and tumor necrosis factor, were found to be perturbed in the same direction, suggesting a shared pharmacological impact of statins and antidepressants on the inflammatory pathways (Supplementary File 5). Widespread enrichment of immune pathways was not observed for genes perturbed in the opposite direction (Supplementary Figure 9).

As control compounds that are not known to have anti-depressive and lipid-lowering effects, alvespimycin displayed limited similarity with statins (average Tau = 23.7), while sirolimus showed an average connectivity score of 92.5. However, unlike antidepressants, genes perturbed in the same direction by statins and sirolimus were not functionally enriched in immune processes, and the high Tau scores between statins and sirolimus appeared to be driven by genes involved in non-lipid metabolic processes, namely nucleobase-containing compound and protein metabolic processes, as well as cellular and biological regulation processes, such as cell cycle regulation and cellular component biogenesis (Supplementary Figure 10), demonstrating the specificity of the pathways commonly perturbed by statins and antidepressants.

### Genetically predicted statin on-target and off-target inhibition is associated with changes in haematological traits

Concordant with previous findings (15–17), MR analysis showed that reduced *HMGCR* expression in blood was specifically associated with reduced LDL-C while showing no association with HDL-C and TG levels, and was associated with decreased CAD risk, increased BMI and T2D risk (Figure 2 and Supplementary File 7), demonstrating the suitability of this approach for investigating the effects of HMGCR inhibition.

**Figure 2.**
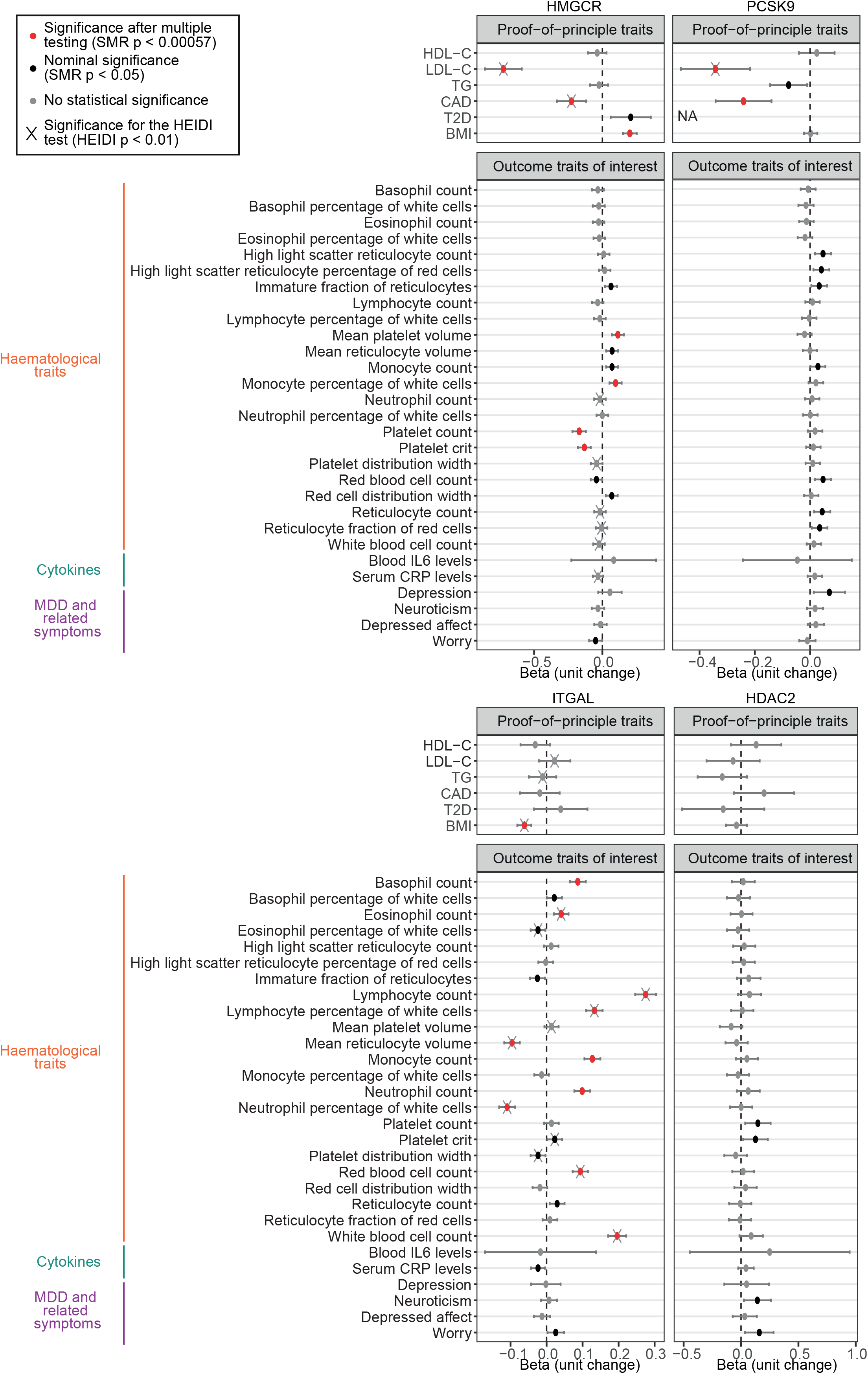
MR analyses of *HMGCR, ITGAL, HDAC2* and *PCSK9* gene expression in blood with various haematological, cytokine and depression-related traits. Dot plots show the associations (beta) between gene expression and traits, and error bars show the 95% confidence intervals. The effect sizes are harmonized to represent the changes in trait per one standard deviation decrease in gene expression. The beta values thus reflect the direction of association upon genetically proxied target inhibition. The units of beta values are not standardized. Red dots represent associations with statistical significance after multiple testing correction (p < 0.00057), and black dots represent associations with nominal significance (p < 0.05). Associations with significant HEIDI p-values are marked by ×. For *PCSK9*, the genetic instrument was not profiled in the type II diabetes (T2D) dataset and the result was thus labelled with “NA” (“not available”).

We did not observe an association of genetically predicted HMGCR inhibition with altered risk of depression, neuroticism or depressed affect; however, we found nominal association with reduced worrying symptoms (*β* = −0.05 (95% CI = −0.097 − −0.0018); p = 0.042) (Figure 2). MR analysis uncovered substantial associations between genetically predicted HMGCR inhibition and changes in haematological traits, including a positive association with mean platelet volume (*β* = 0.11 (95% CI = 0.069 − 0.16); p = 4.49E-07), and a negative association with platelet count (*β* = −0.17 (95% CI = −0.22 − −0.12); p = 1.35E-11) and platelet crit (volume occupied by platelets in the blood as a percentage) (*β* = −0.13 (95% CI = −0.18 − −0.085); p = 3.14E-08) (*β* values are in the unit of standard deviation (SD) change in outcome trait per 1-SD decrease in *HMGCR* gene expression) (Figure 2). In addition, the reduction in *HMGCR* expression showed a significant association with increase in monocyte percentage (*β* = 0.097 (95% CI = 0.052 − 0.14); p = 2.18E-05). We observed no association between *HMGCR* expression and inflammatory cytokine levels, namely IL6 and CRP.

Interestingly, we observed HEIDI significance for the association of *HMGCR* expression in blood with CAD and LDL-C (Figure 2), which suggested that the variant associated with *HMGCR* expression was distinct from those associated with CAD risk and LDL-C level, despite extensive validation of the effects of HMGCR inhibition on these traits by previous studies (18, 19). This is likely due to the complex LD structures in the GWAS studies (Supplementary Figure 11). Sensitivity analysis using *HMGCR* expression in brain showed concordant patterns of associations with haematological and depression-related traits (Supplementary Figure 12 and Supplementary File 8).

In analyses of off-target effects, we observed no association between *ITGAL* and *HDAC2* blood expression with lipid traits, confirming that the lipid-lowering effects of statins were driven by on-target HMGCR inhibition (Figure 2). We found significant associations of genetically proxied ITGAL inhibition with BMI and changes in haematological traits (Figure 2 and Supplementary File 9). Limited associations were found between *HDAC2* expression and outcome traits assessed in this study (Figure 2 and Supplementary File 10). Overall, genetically proxied statin target inhibition was found to be associated with changes in diverse haematological traits through both on-target and off-target effects.

To assess whether the observed associations were specific to statins and not other LDL-C-lowering drugs, we performed SMR analysis to predict the effects of Proprotein Convertase Subtilisin/Kexin type 9 (PCSK9) inhibitors (Figure 2 and Supplementary File 11). As expected, genetically predicted PCSK9 inhibition showed a substantial association with reduced LDL-C and CAD risk, but no significant association with platelet traits, indicating that the latter were likely to be independent of lipid-lowering effects of statins. Interestingly, genetically predicted PCSK9 inhibition was nominally associated with increased depression risk, consistent with previous findings (20).

## Discussion

To our best knowledge, this is the first study that directly compares and explores the commonality in the gene expression responses upon statin and antidepressant exposure, and provides genomic evidence for shared pharmacological mechanisms of the two drug classes. Using *in vitro* models, we found substantial similarities in gene expression changes induced by statin and antidepressant exposure, conferred by concordant impacts on cellular processes, biological regulation and diverse immune pathways, suggesting a shared pharmacological effect of both drug classes. Using MR, we found no genetic association of HMGCR inhibition with depression risk, but observed extensive associations with various haematological traits, particularly monocytes and platelets, further supporting an immunomodulatory activity of statins. Our study design and main findings are summarized in Figure 3.

**Figure 3.**
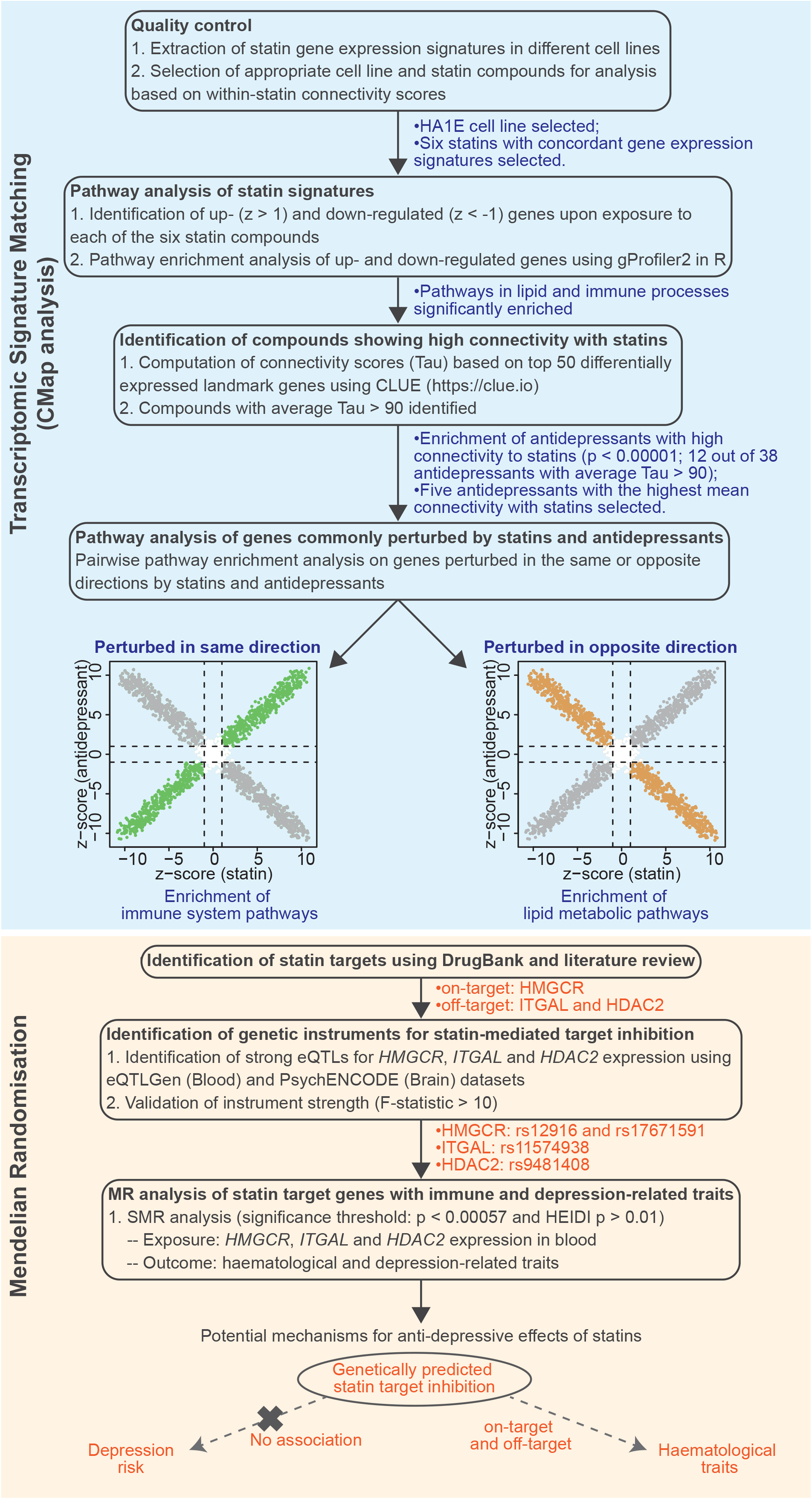
Summary of study design and main findings. We explored the gene expression changes induced by statins using the pre-compiled Connectivity Map (CMap) gene expression signatures, and identified biological processes perturbed by statins and antidepressants in the same or opposite direction. We performed Mendelian randomization (MR) analysis to investigate the association of genetically predicted statin target inhibition with various haematological and depression-related traits.

Corroborating our results, previous MR studies found no association between LDL-C and depression (21, 22). Another MR study reported an association between genetically proxied HMGCR inhibition and increased depression risk; however, given they used multiple genetic instruments in the *HMGCR* gene that are in moderate LD (r^2^ = 0.393 in European population), their reported effect is likely to be inflated (23).

Immune system dysfunction and elevated pro-inflammatory cytokine levels have been linked to disturbed serotonin metabolism and depressive symptoms (24, 25). Furthermore, conventional antidepressants exhibit immunomodulatory properties, by reducing the levels of pro-inflammatory cytokines (24). Though we did not find any association of HMGCR inhibition with IL6 and CRP through MR analysis, we found that pathways related to other pro-inflammatory cytokines, namely interleukin-1 and tumor necrosis factor (TNF), were frequently enriched amongst genes downregulated by statins (Supplementary File 2), corroborating previous findings (6). Furthermore, MR uncovered extensive associations between genetically predicted HMGCR inhibition and alterations in various blood cell parameters, including monocytes and platelets. Depressed individuals have been found to exhibit increased platelet activity (26), with platelet activity reduced after antidepressant treatment (27, 28). Similarly, monocyte activation has been described in depression (29).

Despite strong evidence of a role of the immune response and inflammatory biomarkers in depression, previous clinical trials of treatments for depression that specifically target these pathways have failed (30). This may be due to an underrepresentation of inflammatory phenotypes within case participants-individuals with increased inflammation are estimated to make up less than one-third of the depressive population (30). Furthermore, common assessment measures of depression, such as the 17-item Hamilton Depression Rating Scale, incorporate physical, cognitive, and emotional symptoms (31). Such combined measures may be limited in detecting symptom-specific effects of immunomodulatory drugs, as infliximab, a TNF antagonist, has shown preferential effects on anhedonic symptoms (32). Corroborating these findings, our MR analysis found that genetically predicted HMGCR inhibition was nominally associated with reduced worrying symptoms, while showing no association with the other symptoms assessed. Therefore, better study designs, including stratification of participants based on immune profiles, assessment of relevant inflammatory biomarkers as secondary measures, and assessment of specific symptoms, are required to evaluate the efficacy of drugs with immunomodulatory and anti-inflammatory properties, such as statins, on treating depression.

Statins are reported to exhibit off-target inhibition of ITGAL and HDAC2, although *in vitro* interactions do not necessarily translate to *in vivo* pharmacological actions (33–36). While no association was observed with depression or depressed affect, genetically predicted ITGAL and HDAC2 inhibition was nominally associated with increased worrying and/or increased neuroticism, which was in the opposite direction to HMGCR inhibition. Although *ITGAL* expression is absent in HA1E cells, and therefore unlikely to contribute to the immunomodulatory effects of statins observed in CMap analysis, MR demonstrated that lowering of *ITGAL* expression in blood was associated with extensive immune cell changes. Furthermore, genetically proxied ITGAL inhibition, but not HMGCR inhibition, was associated with reduced CRP levels, suggesting that the CRP-lowering effects of statins could potentially be mediated through off-target pathways. Given its expression and functional involvement in diverse immune cell populations (37), inhibition of ITGAL likely leads to extensive alterations in immune phenomena. Taken together, our findings suggest that statins exhibit modulatory effects on the immune system and possibly depressive symptoms through both on-target HMGCR inhibition, and potentially off-target ITGAL inhibition.

Most previous studies on the association between statins and depression have been observational studies or RCTs. Our study offers a genomics approach to investigating these effects at the molecular level, via transcriptomic signature matching and genetic association analysis. We performed MR analysis using large eQTL and GWAS datasets, which is less prone to unmeasured confounder bias and reverse causality than observational studies.

Several limitations need to be acknowledged. Firstly, the dosage of statins used to generate the CMap gene expression signatures might not reflect the doses used in treatment, and the immortalised cell lines potentially exhibit genomic variations from biological tissues. Secondly, while protein quantitative trait loci (pQTL) provide better genetic instruments for statin-mediated target protein inhibition, we were limited by pQTL data availability, and thus used strong and previously validated eQTLs as instruments for HMGCR inhibition. Additionally, the associations observed for genetic variants provide an estimated effect of lifelong statin exposure and may not reflect short-term effects. However, investigating the long-term effect of statins is clinically relevant, as they are often taken until the end of life (38). Finally, different statin compounds, varying in pharmacokinetic characteristics, display different ability to cross the blood-brain barrier and association with depression (7). A comparative analysis between different statins may provide insights into the chemical properties underlying their anti-depressive effects.

In conclusion, we show that statins evoke an antidepressant-like effect on various immune activation and response processes in human cell lines, and demonstrate modulatory impacts of statins on extensive haematological phenomena through both on-target and off-target effects. At the time of writing, there are ongoing clinical trials investigating the effect of statin treatments on depression and emotional processing (39–42). Our findings provide justification for future clinical trials investigating the effects of statins on depressive symptoms, especially in relation to blood inflammatory biomarkers, which can be used as secondary measures, as well as to stratify participants and identify sub-cohorts that are most likely to respond to statin treatments.

## Data Availability

All data produced in the present work are contained in the manuscript or available upon reasonable request to the authors.

https://github.com/CNSGenomics/statin_depression

## Acknowledgements

JCJ is supported by an NHMRC IDEAS grant (2000637). AMM is supported by the Wellcome Trust (216767/Z/19/Z, 220857/Z/20/Z, 223165/Z/21/Z) and UKRI MRC (MR/W014386/1). SS is supported by funding from the National Health and Medical Research Council (NHMRC) Program Grant (1113400), NHMRC Early Career Fellowship (APP1142495) and National Heart Foundation Future Leader Fellowship (105638).

The publicly available CMap dataset was retrieved from the GEO repository (GSE92742). All GWAS summary statistics analyzed in this study were retrieved from original publications or public platforms, as referenced in the manuscript. Analysis scripts can be found at https://github.com/CNSGenomics/statin_depression. This research has been conducted using the UK Biobank Resource under Application Number 12505.

We thank Professor Naomi Wray for her comments and suggestions for the manuscript. We thank Dr Zhihong Zhu, Dr Zhili Zheng and Dr Yang Wu for their insightful comments on the SMR tool.

