## Supplementary Methods, Figures and Tables for "Statins and antidepressants induce similar *in vitro* gene expression responses"

**of**

### Supplementary Methods

#### Bioinformatic analyses

All R analyses described in this study were performed in R version 4.1.0 unless otherwise stated.

#### CMap gene expression signatures

The Connectivity Map (CMap) database was first introduced in 2006 (1). The Library of Integrated Cellular Signatures (LINCS) database from 2017 represents a 1 000-fold scale-up of the original CMap database and contains a diverse collection of transcriptomic responses to chemical and genetic perturbations, primarily profiled using human cell lines (1, 2). For each perturbagen, its transcriptomic impacts of different treatment dosages and timepoints are investigated using the L1000 platform, which directly measures the gene expression of 978 genes (termed "landmark" genes) (2). The expression levels of 11 350 genes are computationally inferred from the landmark genes, constructing a transcriptomic profile consisting of a total of 12 328 genes (2). The L1000 platform generates gene expression profiles that show high correlation with those measured using RNA-seq, validating its use as a feasible and cost-effective approach for high-throughput profiling of perturbagen transcriptomic profiles (2). In this study, the level 5 z-scores of the CMap signatures, which represent a normalized measure of the magnitude and direction of gene expression changes induced by the perturbagens, were acquired from the GEO repository (GSE92742) in the GCTX format (accessed on July 8$^{th}$, 2021).

#### Selection of statin gene expression signatures

The transcriptomic signatures of statins from the HA1E kidney cell line were selected for detailed analysis, due to the greatest concordance and specificity in gene expression changes amongst different statins (Supplementary Figure 1). Previous validation of the CMap pipeline shows that drugs are able to elicit intended pharmacological responses even in cell lines that are not derived from their primary target tissue types, provided that their targets are expressed in the cell lines (1). We queried the Protein Atlas (3) and DepMap expression platforms (4), and confirmed that 3-Hydroxy-3-Methylglutaryl Coenzyme A Reductase (HMGCR) was ubiquitously expressed in diverse human tissues and cell lines, including the kidney and HA1E cells. Detailed analysis of statin gene expression signatures was performed on signatures generated in selected experimental conditions (perturbation time = 24 h, perturbation dose = 10 μM). The MCF7 (breast cancer cell line) and HepG2 (liver cancer cell line) transcriptomic profiles of statins were also analyzed for the purpose of comparison. The gene expression signatures of alvespimycin (heat shock protein inhibitor) (5) and sirolimus (mTOR inhibitor) (6) were analyzed as controls, as they were reported by the CMap team to induce consistent and strong transcriptomic responses across cell lines, and are not functionally linked to lipid-associated pathways or strong anti-depressive effects.

**CMap connectivity scores**

The CMap platform uses an algorithm based on the weighted Kolmogorov-Smirnov enrichment statistic to quantify the similarity between two signatures (2). The degree of connectivity is computed as Tau scores, which represent a statistical measure of the likelihood of observing the similarity given all transcriptomic signatures in the reference database. The Tau scores range between -100 and 100, where a positive Tau score indicates positive connectivity. The pre-compiled Tau scores between reference signatures in the CMap database, which are available on the online CLUE platform (https://clue.io), are computed using the top 50 up-regulated landmark genes and the top 50 down-regulated landmark genes.

**Transcriptome-wide pairwise correlation**

We further interrogated the gene expression correlation of all 12 328 genes profiled in each CMap signature. Pairwise correlation between a pair of signatures was evaluated using a simple linear regression model in R. The regression coefficient (slope), p-value, and adjusted $R^2$ value for each linear regression model were summarized.

**Annotation of biological process terms with ancestor terms**

Gene Ontology (GO) biological process terms are placed in a hierarchical structure consisting of parent/child term relationships (7). While functional annotation by gProfiler2 provides detailed information on the enriched biological processes, the resulting list of significant terms often contains overlapping and highly redundant biological processes, confounding the interpretation of results. We thus summarized the GO biological process terms based on their high-level ancestor terms. Using GO.db (version 3.13.0) (8), we retrieved the child terms of biological process (GO:0008150), which is the root term of all GO biological pathway terms, and defined these child terms as the list of ancestor biological process terms used for downstream annotation. We annotated each statistically significant GO biological process term based on their ancestor categories. Each GO term might be annotated with more than one ancestor category. Similarly, we also categorized the GO biological terms into primary metabolic process (GO:0044238) (a child term of "Metabolic process") and immune system process (GO:0002376) terms.

**Investigating gene expression changes related to GO-annotated biological pathways**

To further interrogate gene expression changes related to specific pathways, z-scores of genes annotated by GO as being functionally associated with lipid pathways were profiled. Briefly, the gene sets for the "lipid metabolic pathway" (GO:0006629) term were obtained from gProfiler2 and converted to Entrez gene IDs using gConvert in R (9, 10). For each gene set, the z-scores of genes profiled by the LINCS dataset were retrieved and visualized on scatterplots.

**Annotating antidepressants**

The Anatomical Therapeutic Chemical (ATC) classification system provides a reference for drugs, which are hierarchically categorized based on their targeted organs as well as their therapeutic, pharmacological and chemical attributes (11). While we acknowledge that there are non-ATC-profiled drugs or active substances with anti-depressive properties, in this study we limited our analysis to antidepressants documented by the ATC system (ATC code: N06A) as they were more likely to have established efficacy and thereby provided a valid basis for interrogating the molecular mechanisms underlying their anti-depressive effects. A total of 38 ATC-documented antidepressants are profiled in HA1E cells by the CMap database.

**Identifying shared perturbed pathways between statins and antidepressants**

We hypothesized that if statins and antidepressants indeed exhibited shared pharmacological effects, they would likely perturb the same biological pathways. Out of the 38 antidepressants profiled in HA1E cells, we selected five antidepressants (desipramine, nortriptyline, paroxetine, sertraline and trimipramine) for further analysis as they demonstrated the highest average connectivity with the six statins studied. We performed pairwise comparison of the transcriptomic signatures of six statins against the five antidepressants, which gave rise to a total of 30 statin-antidepressant combinations. For each pair of statin and antidepressant, genes that were differentially expressed (defined by $|z| > 1$) in both signatures were further categorized into genes perturbed in the same or opposite direction. Using gProfiler2 and parameters described in Methods, pathway enrichment analysis was performed for each category. Similarly, we also performed pairwise pathway enrichment analysis for genes commonly perturbed by statins and the two control drugs (alvespimycin and sirolimus).

**Mendelian randomization**

The principle underlying Mendelian randomization (MR) originates from Mendel's laws of segregation and independent assortment. By Mendel's laws, alleles randomly segregate and are passed from parents to offspring during the process of meiosis, and the inheritance of alleles of one variant occurs independently of alleles of other variants (12). In randomized controlled trials, participants are randomly allocated to the treatment or control groups, and thereby subjected to different levels of exposure. MR uses genetic instruments linked to the exposure of interest as proxies, as if the level of exposure is randomly allocated at conception. This random assignment of genetically determined exposure at conception is less likely to be affected by various lifestyle and socioeconomic factors, which may otherwise distort the exposure-outcome associations (13, 14). Furthermore, as the genetic determinants for exposures are generally fixed at conception and thus not affected by the outcome, MR is less prone to reverse causation, which may be problematic to dissect using observational studies (15). Genetic variants are only valid instrument variables for MR analysis if they meet three key assumptions, that they are strongly associated with the exposure, not associated with any confounder of the exposure-outcome association, and are associated with the outcome only through the exposure of interest (no horizontal pleiotropy) (16).

The selection of genetic instruments depends on the genetic architecture of the exposure (17). For polygenic traits, such as low-density lipoprotein cholesterol (LDL-C) levels, the genetic proxy for these traits is derived from genetic variants from multiple trait-associated loci (17). In contrast, in cases where the exposure is a protein, such as HMGCR, its genetic instruments may be selected from expression quantitative trait loci (eQTL) or protein quantitative trait loci (pQTL) located within or nearby the protein-encoding gene (17). Previous studies have reported an enrichment of eQTLs in single nucleotide polymorphisms (SNP) associated with complex traits, and suggested gene expression as an important mediator of SNP-trait associations, supporting the utility of eQTLs in identifying genetic determinants of traits and diseases (18, 19). Furthermore, eQTLs are widely used as genetic proxies for drug targets in MR analyses (20, 21).

In this study, we assessed the strength of genetic instruments using the F-statistics from the linear regression model of the genetic instruments with the exposure (gene expression). We determined SNPs with F-statistic > 10 to be suitable genetic instruments, where F-statistic was calculated as previously described (17, 20).

**eQTLGen blood eQTLs**

The eQTLGen consortium has produced an extensive catalogue of both *cis* and *trans* eQTLs for human gene expression in blood (22). The blood eQTL data were generated via meta-analysis of over 31 684 samples from 37 cohorts consisting of primarily European individuals (22). In the current study, only cis-eQTLs were considered for the selection of genetic instruments.

**GTEx tissue eQTLs**

The GTEx project (version 8) identified and profiled eQTLs from diverse tissue types (23). The majority of the GTEx cohort consists of individuals of European descent. We retrieved Proprotein Convertase Subtilisin/Kexin type 9 (*PCSK9*) eQTLs identified in whole blood, which contains 670 samples.

**PsychENCODE eQTLs**

To use genetic instruments relevant to the biology of depression, we retrieved brain eQTL data produced by the PsychENCODE consortium. The PsychENCODE eQTL dataset contains prefrontal cortex eQTLs identified in a cohort of 1 387 individuals (24).

**Genetic instrument for HMGCR inhibition**

To investigate the on-target effect of statins, we selected rs12916 as the genetic proxy for HMGCR inhibition. The rs12916 SNP is located in the 3′ untranslated region of the *HMGCR* gene, and the genetic associations between the rs12916-T allele and metabolomic traits were shown to mirror the actual effects of statin exposure on the metabolome (25). It is widely used as a genetic proxy for assessing statin effects (26-28). Rs12916 is a strong eQTL for *HMGCR* expression, where each additional rs12916-T allele is associated with 0.1 standard deviation (SD) decrease in *HMGCR* expression in blood (eQTLGen: p = 1.5E-36). As a sensitivity analysis, we also selected rs17671591, a strong eQTL (p = 2.5E-05) for *HMGCR* expression in the brain prefrontal cortex (PsychENCODE (24)), as the genetic instrument for HMGCR inhibition in the brain. Rs17671591 is in moderate linkage disequilibrium (LD) ($r^2 = 0.6$) with rs12916 amongst European individuals.

**Statin off-target inhibition of ITGAL and HDAC2**

Integrin Alpha-L (ITGAL) is a component of the heterodimeric Lymphocyte Function-associated Antigen-1 (LFA-1) receptor, which is expressed in diverse immune cell populations and plays a role in modulating cell adhesion (29, 30). Lovastatin and simvastatin are found to show *in vitro* binding to ITGAL, subsequently inhibiting LFA-1 function and T-cell adhesion, proliferation, and cytokine production (31-33). At the same time, a subset of statins (atorvastatin, fluvastatin, lovastatin, pravastatin and simvastatin) are also found to exhibit *in vitro* inhibition of Histone Deacetylase 2 (HDAC2) (34), which is involved in chromatin remodelling (35).

Using the eQTLGen data (22), we selected the most significant eQTLs in blood to proxy for inhibition of ITGAL and HDAC2. Specifically, each additional rs11574938-C was associated with 0.21 SD decrease of *ITGAL* expression ($p = 7.9E-150$), and each rs9481408-T allele was associated with 0.048 SD decrease of *HDAC2* expression ($p = 4.1E-07$) in blood.

**Genetic instrument for PCSK9 inhibition**

We sought to investigate whether any genetic associations observed for statin targets were likely to be mediated independently of cholesterol lowering, by extending MR analysis to other lipid-lowering medications, namely PCSK9 inhibitors (36). *PCSK9* eQTLs were absent in the eQTLGen dataset, as *PCSK9* was likely removed from meta-analysis due to the lack of variation in expression. We thus identified *PCSK9* eQTLs from the GTEx whole blood dataset, and performed MR analysis using a strong eQTL (rs12117661; $p = 8.31E-11$) as the genetic instrument for *PCSK9* expression.

**HEIDI test**

To validate that the exposure-outcome association was mediated through one single causal SNP, rather than via LD between separate SNPs (linkage scenario), the heterogeneity in dependent instruments (HEIDI) test was performed as a sensitivity analysis (37). Without correcting for multiple testing, we used a HEIDI p-value threshold of 0.01 to define statistical significance, which was a stringent threshold. A HEIDI p-value > 0.01 indicated that the estimated effect of exposure on outcome was likely due to single causal variants, while a p-value < 0.01 suggested that the observed association was potentially attributable to the linkage scenario.

**LocusCompare plots**

LocusCompare plots allow the visualization of the distribution of genome-wide association studies (GWAS) or eQTL summary statistics (38). Using the LocusCompareR package (version 1.0.0) (38), we generated the LocusZoom and significance ($-\log_{10}P$) scatter plots for LDL-C and *HMGCR* eQTLs from eQTLGen. The LD scores were generated based on the European population.

### Supplementary Tables

**Supplementary Table 1.** The CMap signatures selected for analyses

| Compound | Cell line | CMap signature |
|---|---|---|
| Atorvastatin | HA1E | CPC006_HA1E_24H:BRD-U88459701-000-01-8:10 |
| Fluvastatin | HA1E | CPC004_HA1E_24H:BRD-K66296774-001-02-0:10 |
| Lovastatin | HA1E | CPC003_HA1E_24H:BRD-K09416995-001-21-7:10 |
| Mevastatin | HA1E | CPC001_HA1E_24H:BRD-K94441233-001-03-1:10 |
| Pravastatin | HA1E | CPC002_HA1E_24H:BRD-K60511616-236-01-4:10 |
| Rosuvastatin | HA1E | CPC004_HA1E_24H:BRD-K82941592-238-02-9:10 |
| Simvastatin | HA1E | CPC005_HA1E_24H:BRD-A81772229-001-01-6:10 |
| Desipramine | HA1E | CPC004_HA1E_24H:BRD-K60762818-003-15-3:10 |
| Nortriptyline | HA1E | CPC002_HA1E_24H:BRD-K91263825-001-03-6:10 |
| Paroxetine | HA1E | CPC004_HA1E_24H:BRD-K37991163-003-06-8:10 |
| Sertraline | HA1E | CPC004_HA1E_24H:BRD-K82036761-003-07-0:10 |
| Trimipramine | HA1E | CPC004_HA1E_24H:BRD-A19195498-050-09-1:10 |
| Alvespimycin | HA1E | CPC014_HA1E_6H:BRD-K83988098-003-01-8:10 |
| Sirolimus | HA1E | CPC012_HA1E_6H:BRD-K89626439-001-01-0:10 |
| Atorvastatin | HepG2 | CPC006_HEPG2_6H:BRD-U88459701-000-01-8:10 |
| Fluvastatin | HepG2 | CPC004_HEPG2_6H:BRD-K66296774-001-02-0:10 |
| Lovastatin | HepG2 | CPC009_HEPG2_6H:BRD-K09416995-001-31-6:10 |
| Mevastatin | HepG2 | CPC016_HEPG2_6H:BRD-K94441233-001-03-1:10 |
| Rosuvastatin | HepG2 | CPC004_HEPG2_6H:BRD-K82941592-238-02-9:10 |
| Atorvastatin | MCF7 | CPC006_MCF7_24H:BRD-U88459701-000-01-8:10 |
| Fluvastatin | MCF7 | CPC015_MCF7_24H:BRD-K66296774-001-02-0:10 |
| Lovastatin | MCF7 | CPC009_MCF7_24H:BRD-K09416995-001-31-6:10 |
| Mevastatin | MCF7 | CPC011_MCF7_24H:BRD-K94441233-001-09-8:10 |
| Rosuvastatin | MCF7 | CPC015_MCF7_24H:BRD-K82941592-238-02-9:10 |
| Simvastatin | MCF7 | CPC005_MCF7_24H:BRD-A81772229-001-01-6:10 |

**Supplementary Table 2.** Number of up-regulated and down-regulated genes upon statin exposure in HA1E cells, defined using different z-score thresholds

| Statin | Z > 1 | Z < -1 | Z > 1.5 | Z < -1.5 | Z > 2 | Z < -2 |
|---|---|---|---|---|---|---|
| Atorvastatin | 1597 | 1606 | 677 | 685 | 224 | 297 |
| Fluvastatin | 4252 | 1599 | 2431 | 919 | 1017 | 520 |
| Lovastatin | 2508 | 1777 | 1134 | 965 | 457 | 515 |
| Mevastatin | 4750 | 1874 | 2890 | 1141 | 1435 | 697 |
| Rosuvastatin | 2034 | 1502 | 851 | 723 | 318 | 345 |
| Simvastatin | 2701 | 1949 | 1436 | 1129 | 664 | 658 |

**Supplementary Table 3.** Antidepressants profiled in the CMap database

| ATC class | ATC code | Drug name | CMap ID |
|---|---|---|---|
| Non-selective monoamine reuptake inhibitors (N06AA) (tricyclic antidepressant (TCA)) | N06AA01 | Desipramine | BRD-K60762818 |
| | N06AA02 | Imipramine | BRD-K38436528 |
| | N06AA04 | Clomipramine | BRD-K52989797 |
| | N06AA06 | Trimipramine | BRD-A19195498 |
| | N06AA07 | Lofepramine | BRD-K82147103 |
| | N06AA08 | Dibenzepin | BRD-K79145749 |
| | N06AA09 | Amitriptyline | BRD-K53737926 |
| | N06AA10 | Nortriptyline | BRD-K91263825 |
| | N06AA11 | Protriptyline | BRD-K42098891 |
| | N06AA12 | Doxepin | BRD-K36616567 |
| | N06AA16 | Dosulepin | BRD-K54759182 |
| | N06AA17 | Amoxapine | BRD-K02265150 |
| | N06AA21 | Maprotiline | BRD-K03319035 |
| Selective serotonin reuptake inhibitors (N06AB) | N06AB03 | Fluoxetine | BRD-A31159102 |
| | N06AB04 | Citalopram | BRD-A47598013 |
| | N06AB05 | Paroxetine | BRD-K37991163 |
| | N06AB06 | Sertraline | BRD-K82036761 |
| | N06AB07 | Alaproclate | BRD-A14966924 |
| | N06AB08 | Fluvoxamine | BRD-K72676686 |
| | N06AB10 | Escitalopram | BRD-K70301876 |
| Monoamine oxidase inhibitors, non-selective (N06AF) | N06AF01 | Isocarboxazid | BRD-K93332168 |
| | N06AF02 | Nialamide | BRD-K12102668 |
| | N06AF03 | Phenelzine | BRD-K87024524 |
| | N06AF04 | Tranylcypromine | BRD-A43974575 |
| Monoamine oxidase A inhibitors (N06AG) | N06AG02 | Moclobemide | BRD-K07237224 |
| Other antidepressants (N06AX) | N06AX03 | Mianserin | BRD-A19661776 |
| | N06AX04 | Nomifensine | BRD-A29644307 |
| | N06AX05 | Trazodone | BRD-K70778732 |
| | N06AX06 | Nefazodone | BRD-K90789829 |
| | N06AX07 | Minaprine | BRD-K02867583 |
| | N06AX08 | Bifemelane | BRD-K18779551 |
| | N06AX11 | Mirtazapine | BRD-A64977602 |
| | N06AX12 | Bupropion | BRD-A05186015 |
| | N06AX14 | Tianeptine | BRD-A53077924 |
| | N06AX16 | Venlafaxine | BRD-A51714012 |
| | N06AX17 | Milnacipran | BRD-K02227374 |
| | N06AX18 | Reboxetine | BRD-A43974499 |
| | N06AX21 | Duloxetine | BRD-K71103788 |

**Supplementary Table 4.** Sources of eQTL datasets

| Dataset | Tissue (used in the study) | Sample size | Ancestry | Reference |
|---|---|---|---|---|
| eQTLGen | Blood | 31684 | Predominantly European | Võsa, Claringbould (22) |
| GTEx (version 8) | Whole blood | 670 | Predominantly European | Aguet, Anand (23) |
| PsychENCODE | Brain prefrontal cortex | 1387 | Predominantly European | Gandal, Zhang (24) |

**Supplementary Table 5.** Sources of GWAS summary statistics

| Trait and sample size | Source | Access date |
|---|---|---|
| Haematological parameters (408112)<br>• Basophil count<br>• Basophil percentage of white cells<br>• Eosinophil count<br>• Eosinophil percentage of white cells<br>• High light scatter reticulocyte count<br>• High light scatter reticulocyte percentage of red cells<br>• Immature fraction of reticulocytes<br>• Lymphocyte count<br>• Lymphocyte percentage of white cells<br>• Mean platelet volume<br>• Mean reticulocyte volume<br>• Monocyte count<br>• Monocyte percentage of white cells<br>• Neutrophil count<br>• Neutrophil percentage of white cells<br>• Platelet count<br>• Platelet crit<br>• Platelet distribution width<br>• Red blood cell count<br>• Red cell distribution width<br>• Reticulocyte count<br>• Reticulocyte fraction of red cells<br>• White blood cell count | Vuckovic, Bao (39) | May 10th, 2022 |
| Blood IL6 level (8189) | Ahola-Olli, Würtz (40) | September 14th, 2021 |
| Serum CRP level (418642) | Han, Ong (41)<br>(GWAS catalogue ID: GCST009777) | September 21st, 2021 |
| Lipids<br>• HDL-C (187167)<br>• LDL-C (173082)<br>• TG (177861) | Willer, Schmidt (42) | September 6th, 2021 |
| CAD (34541 cases and 261984 controls from the UK Biobank cohort) | van der Harst and Verweij (43)<br>(openGWAS ID: ebi-a-GCST005194) | September 9th, 2021 |
| T2D (62892 cases and 596424 controls) | Xue, Wu (44)<br>(openGWAS ID: ebi-a-GCST006867) | September 9th, 2021 |
| BMI (461460) | Hemani, Zheng (45)<br>(openGWAS ID: ukb-b-19953) | September 10th, 2021 |
| Depression (170756 cases and 329443 controls) (excluding 23andMe data) | Howard, Adams (46) | February 15th, 2022 |
| Neuroticism (380060) | Nagel, Watanabe (47) | September 22nd, 2021 |
| Depressed affect (357957) | Nagel, Jansen (48) | September 22nd, 2021 |
| Worrying (348219) | Nagel, Jansen (48) | September 22nd, 2021 |

**Supplementary Table 6.** Counts of commonly perturbed genes by statins and antidepressants

| Statin | Antidepressant | Same direction | Opposite direction |
|---|---|---|---|
| Atorvastatin | Desipramine | 483 | 79 |
| Fluvastatin | Desipramine | 566 | 165 |
| Lovastatin | Desipramine | 501 | 168 |
| Mevastatin | Desipramine | 653 | 208 |
| Rosuvastatin | Desipramine | 476 | 124 |
| Simvastatin | Desipramine | 565 | 164 |
| Atorvastatin | Nortriptyline | 420 | 66 |
| Fluvastatin | Nortriptyline | 548 | 108 |
| Lovastatin | Nortriptyline | 570 | 90 |
| Mevastatin | Nortriptyline | 609 | 102 |
| Rosuvastatin | Nortriptyline | 458 | 93 |
| Simvastatin | Nortriptyline | 569 | 83 |
| Atorvastatin | Paroxetine | 626 | 112 |
| Fluvastatin | Paroxetine | 1144 | 135 |
| Lovastatin | Paroxetine | 886 | 169 |
| Mevastatin | Paroxetine | 1211 | 166 |
| Rosuvastatin | Paroxetine | 719 | 138 |
| Simvastatin | Paroxetine | 958 | 148 |
| Atorvastatin | Sertraline | 948 | 66 |
| Fluvastatin | Sertraline | 1173 | 192 |
| Lovastatin | Sertraline | 1082 | 157 |
| Mevastatin | Sertraline | 1229 | 241 |
| Rosuvastatin | Sertraline | 987 | 95 |
| Simvastatin | Sertraline | 1250 | 111 |
| Atorvastatin | Trimipramine | 553 | 58 |
| Fluvastatin | Trimipramine | 560 | 753 |
| Lovastatin | Trimipramine | 547 | 233 |
| Mevastatin | Trimipramine | 613 | 834 |
| Rosuvastatin | Trimipramine | 591 | 136 |
| Simvastatin | Trimipramine | 671 | 162 |

### Supplementary Figures

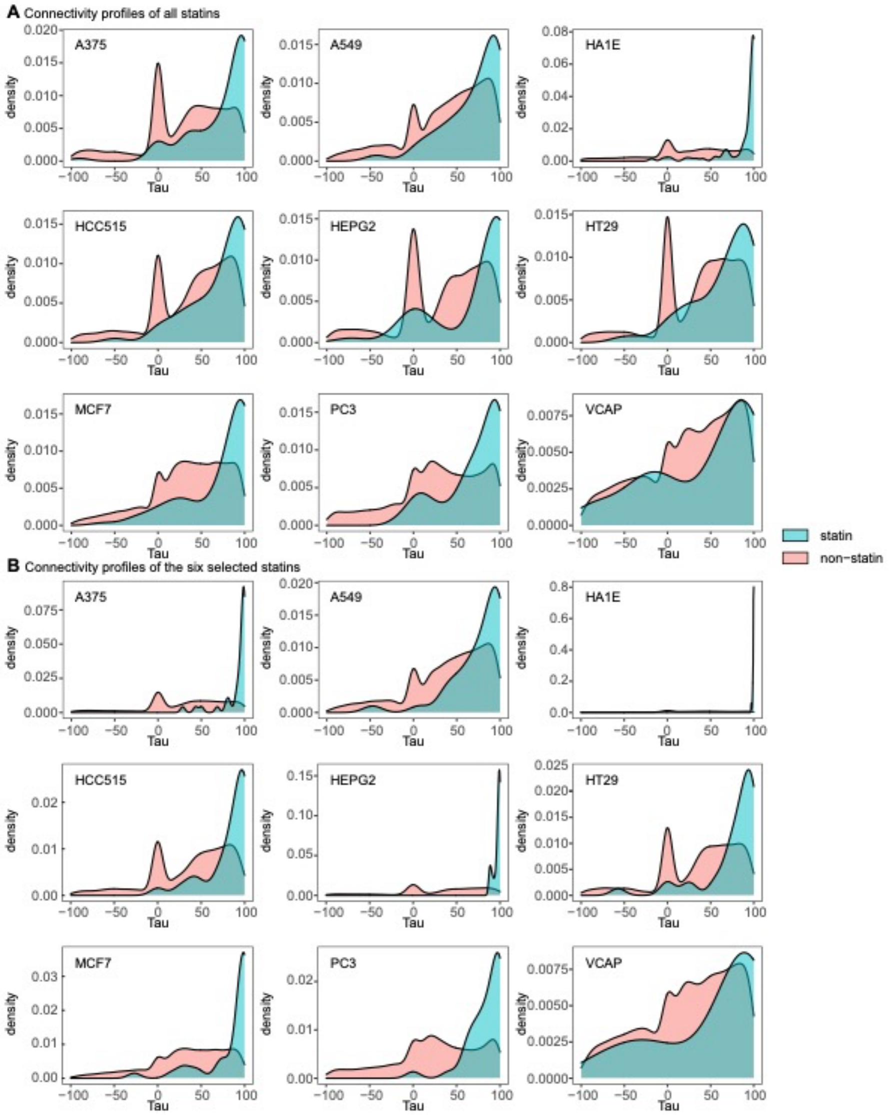

**Supplementary Figure 1.** Kernel density plots of connectivity (Tau) scores computed between different statin compounds (blue), as well as between statins and non-statin compounds (pink) in various cell lines. (**A**) Tau score distribution for all statins (a total of 9, not measured in every cell line) profiled in the CMap database. Compared to the other cell lines, statin-induced gene expression signatures displayed the most concordance in HA1E cells reflected by overall high between-statin connectivity scores (blue), and high specificity, reflected by low connectivity with most other compounds (pink). (**B**) Tau score distribution for six statins (atorvastatin, fluvastatin, lovastatin, mevastatin, rosuvastatin and simvastatin) included in detailed analysis, which were chosen as they were profiled in HA1E cells under selected perturbational conditions (time = 24 h and dose = 10 μM). Pravastatin was not included in analysis as it showed poor connectivity with other statin compounds. The connectivity scores shown in these plots were obtained from the Touchstone dataset, pre-compiled by CLUE (https://clue.io).

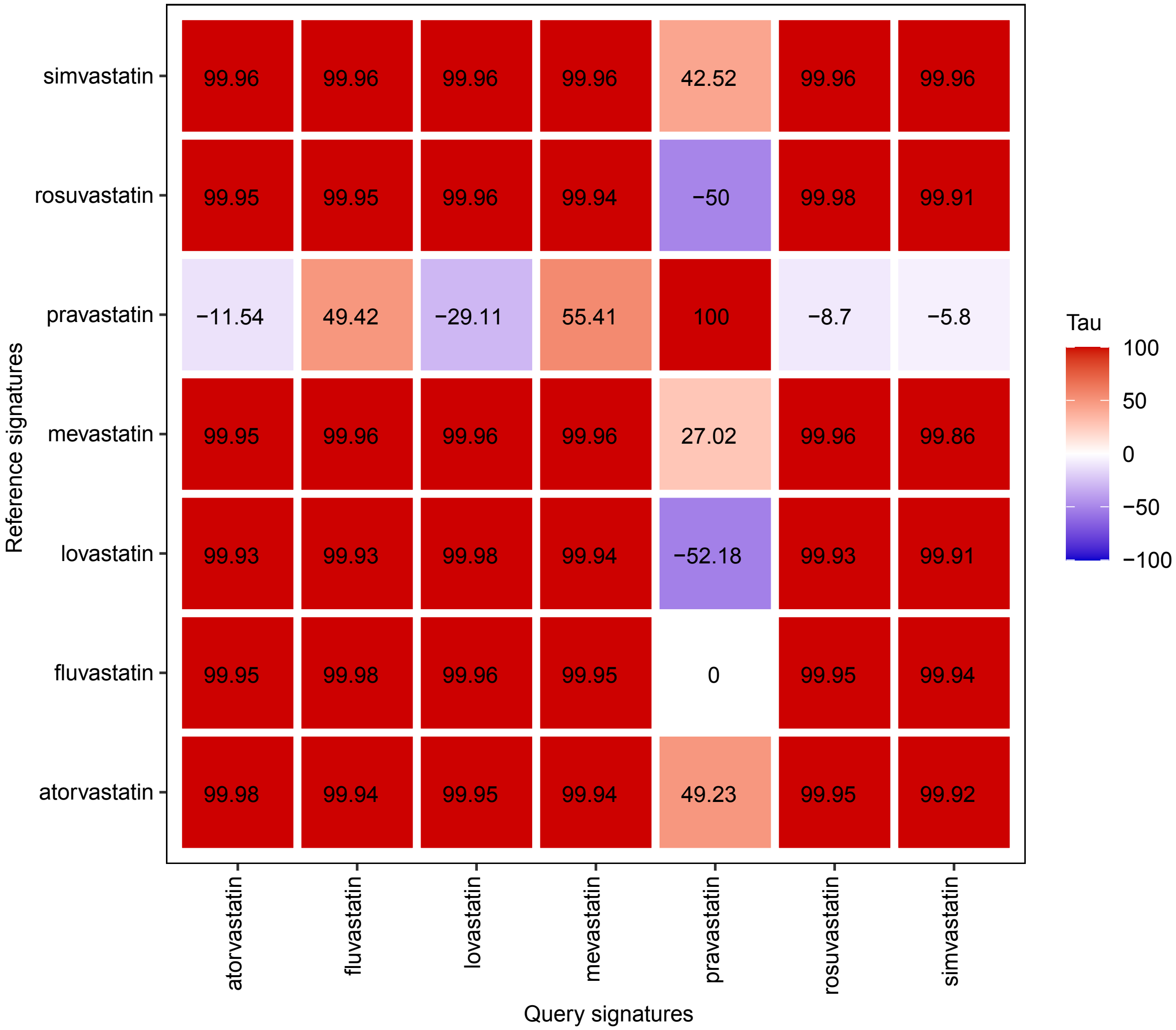

**Supplementary Figure 2.** Connectivity scores amongst statin signatures in HA1E cells. The connectivity scores shown here were generated by submitting statin HA1E query signatures to CLUE (https://clue.io). Columns show the statin query signatures, and rows show the reference statin signatures in the CMap database. The corresponding connectivity scores are annotated on the heatmap.

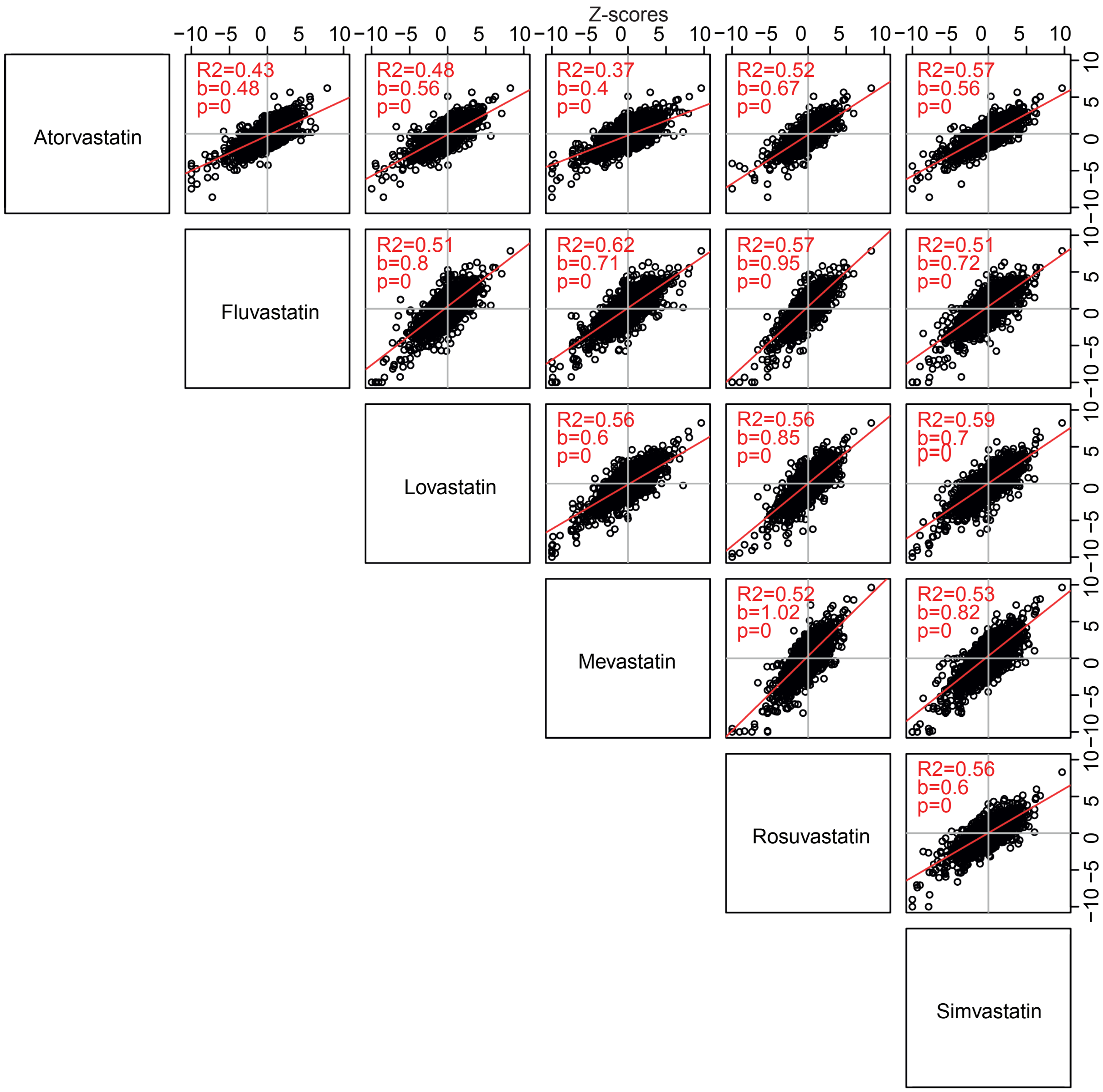

**Supplementary Figure 3.** Statins induced concordant transcriptional responses in HA1E cells. Changes in gene expression (z-scores) of 12 328 genes profiled by CMap were compared for each pair of statins using pairwise correlation. The adjusted $R^2$, the regression coefficients (b) and the corresponding p-values (p) are shown.

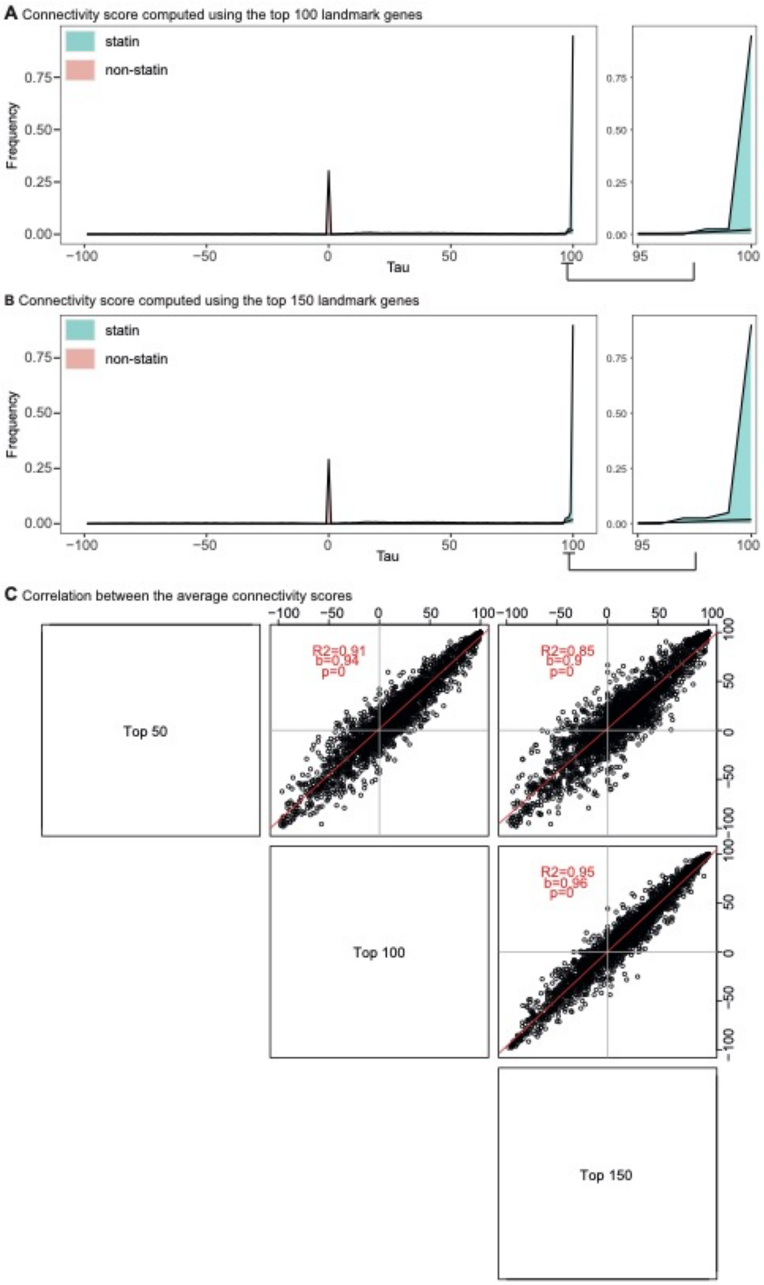

**Supplementary Figure 4.** Statin connectivity profiles computed using different numbers of landmark genes. Connectivity scores were computed using (**A**) the top 100 or (**B**) the top 150 most differentially expressed landmark genes. Distribution (bin = 1 Tau score change) of connectivity scores computed between different statins (blue), as well as between statins and non-statin compounds (pink) are shown. A zoom-in view of connectivity scores between 95 and 100 is shown. (**C**) Pairwise correlation of average connectivity scores (averaged across six statins) between the connectivity profiles computed using the top 50, 100 and 150 most differentially expressed landmark genes. Each dot represents the average connectivity scores of a compound profiled in HA1E cells (total = 2 783).

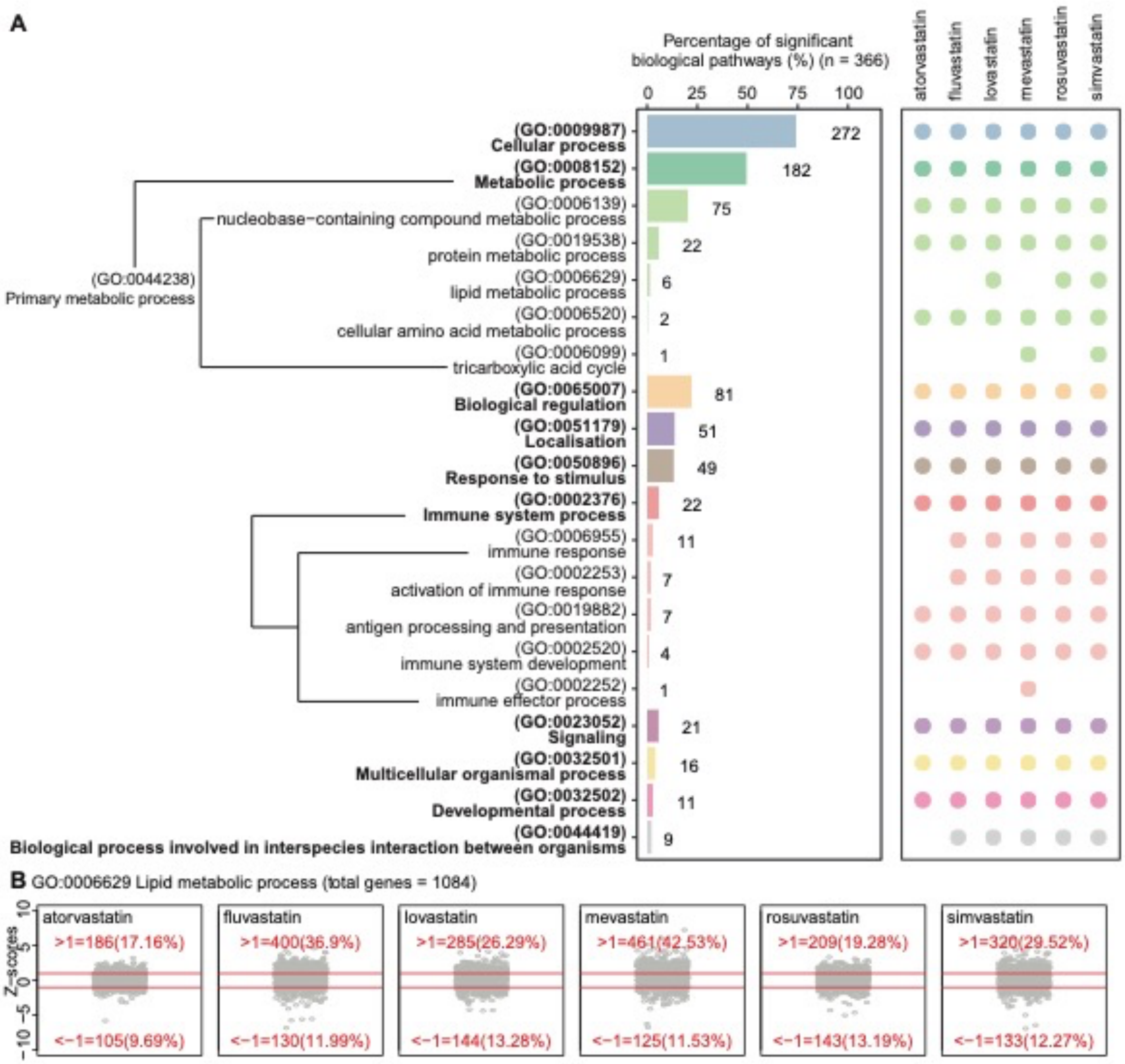

**Supplementary Figure 5.** Genes functionally involved in lipid and immune pathways were widely perturbed by statin exposure in HA1E cells. (**A**) Biological processes identified as significantly enriched amongst statin-induced differentially expressed genes. A total of 366 biological process terms were identified as significant, and categorized into high-level ancestor terms. Y-axis shows the names and GO accession numbers of the ancestor biological process terms, as well as the child terms of primary metabolic process (GO:0044238) (a child term of "metabolic process") and immune system process (GO:0002376). The branches indicate ancestor-child relationships of primary metabolic process and immune system process terms. Bar graph shows the percentages of significant GO biological process terms annotated with each ancestor category, with the corresponding counts shown on the graph. The bubble plot shows the statin compounds for which the biological processes were identified as significantly enriched. (**B**) Statin-induced expression changes of genes functionally involved in the lipid metabolic process (GO: 0006629). Red lines indicate z-scores of 1 and -1, which were used to define up-regulated and down-regulated genes respectively. The total number of genes annotated with the GO term, as well as the numbers and percentages of up-regulated and down-regulated genes are shown.

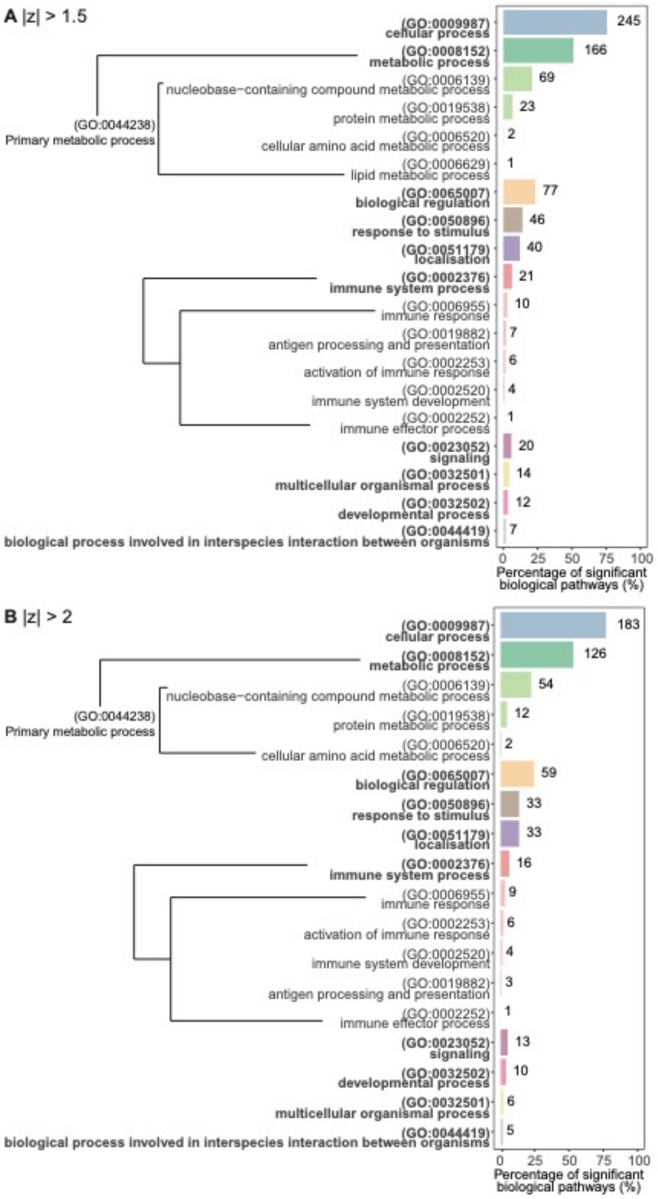

**Supplementary Figure 6.** Pathway enrichment results using more stringent z-score thresholds for defining differentially expressed genes. Pathway enrichment analysis was performed for differentially expressed genes, defined by (**A**) absolute z-score > 1.5 and (**B**) absolute z-score > 2, where a total of 324 and 237 biological pathways were identified as significantly enriched respectively. Enriched biological process terms were categorized into high-level ancestor terms. Y-axis shows the names and GO accession numbers of the ancestor biological process terms, as well as the child terms of primary metabolic process (a child term of "metabolic process") and immune system process. The branches indicate ancestor-child relationships of primary metabolic process and immune system process terms. Bar graph shows the percentages of significant GO biological process terms annotated with each ancestor term, with the corresponding counts shown on the graph.

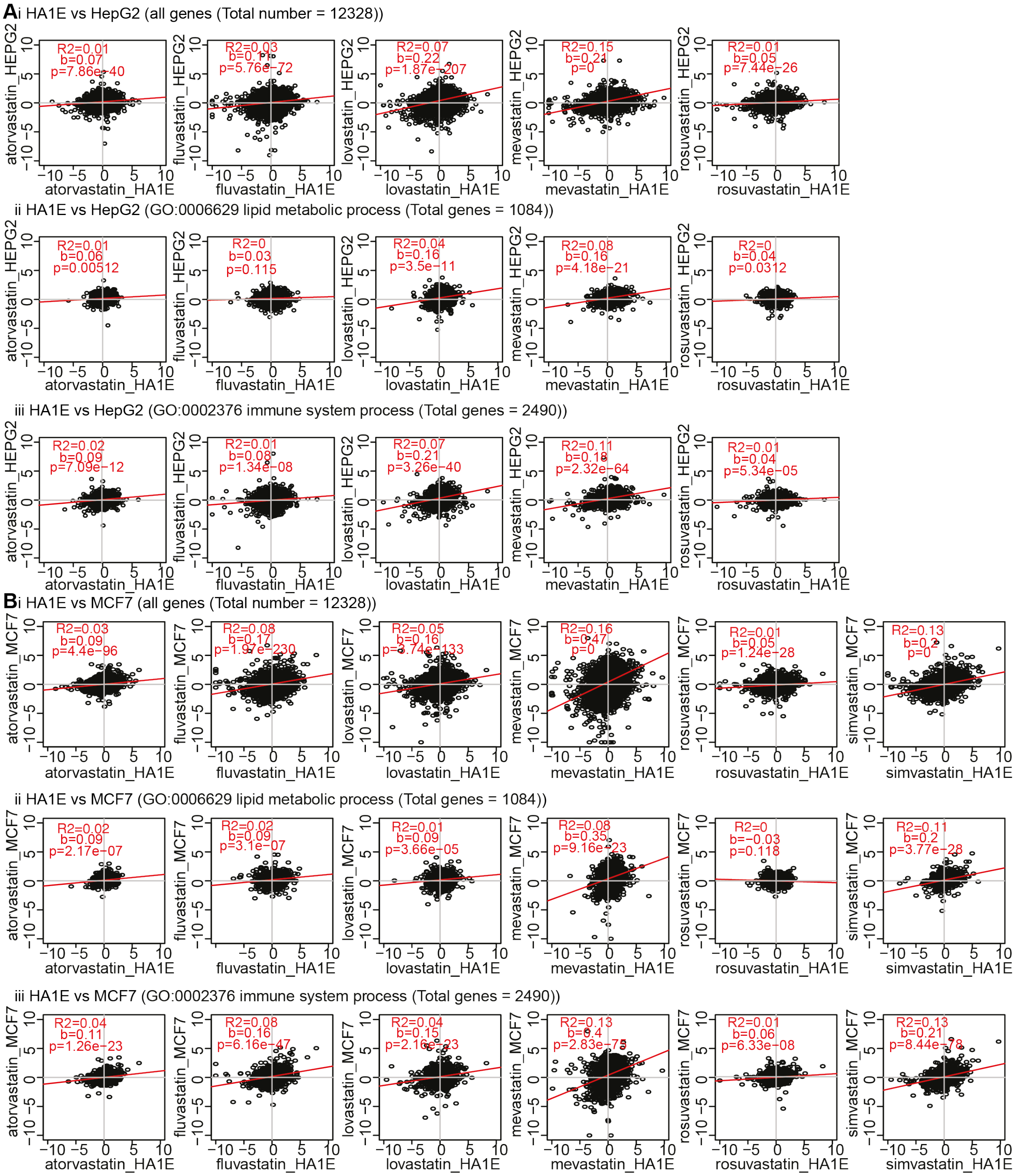

**Supplementary Figure 7.** Pairwise correlation of gene expression changes induced by statins in different cell lines. The correlations between HA1E and (**A**) HepG2 or (**B**) MCF7 cells are shown for the gene expression changes (z-scores) of (i) all 12 328 genes, (ii) lipid-associated genes and (iii) immune-associated genes. The adjusted $R^2$, the regression coefficients (b) and the corresponding p-values (p) are shown.

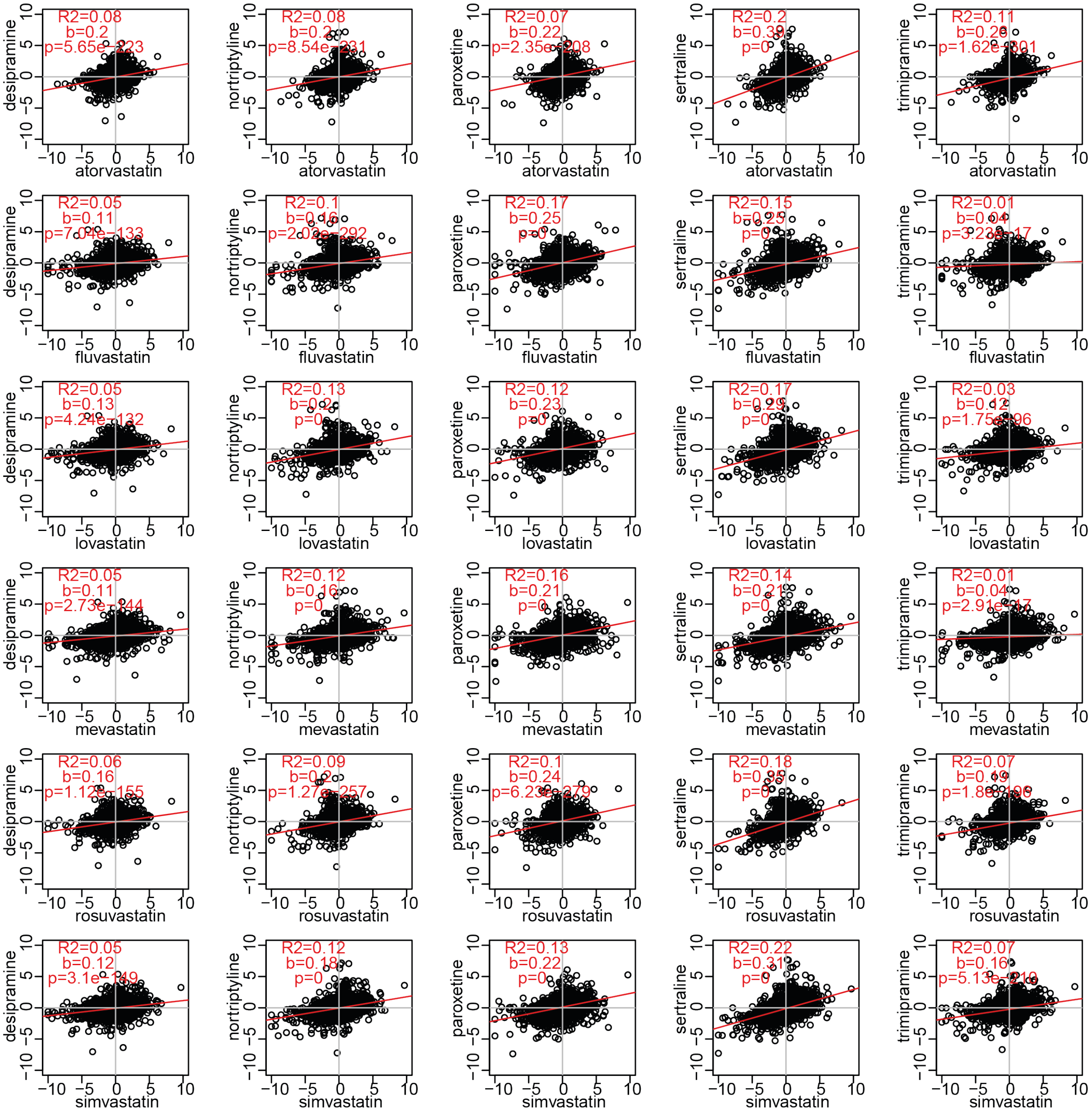

**Supplementary Figure 8.** Pairwise correlation of gene expression changes induced by statins and antidepressants in HA1E cells. The changes in gene expression (z-scores) of 12 328 genes profiled by CMap were compared for each pair of statin and antidepressant. The adjusted $R^2$, the regression coefficients (b) and the corresponding p-values (p) are shown.

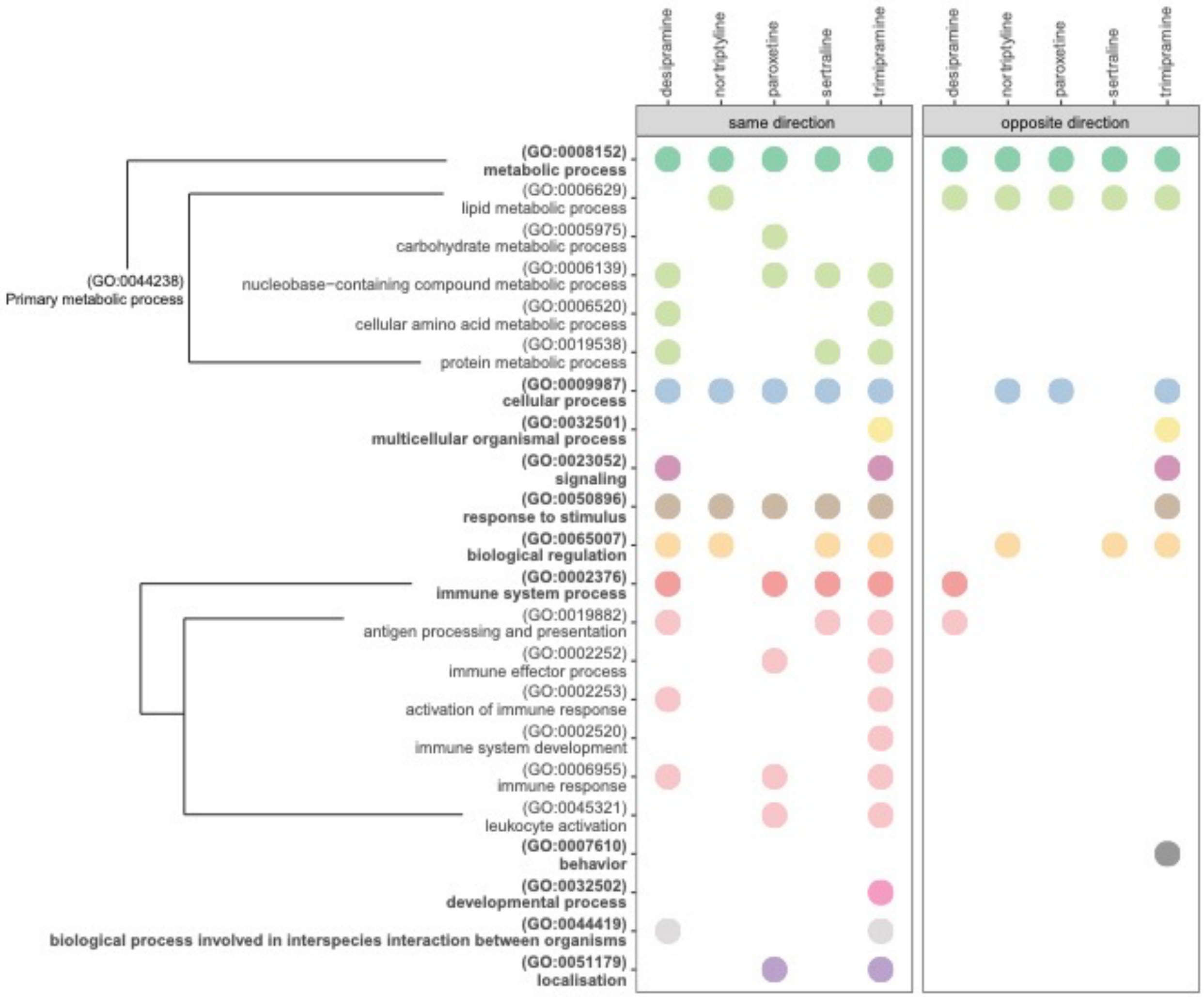

**Supplementary Figure 9.** Biological processes identified as significantly enriched amongst genes perturbed by both statins and antidepressants, in the same (left panel) and opposite (right panel) directions. Y-axis shows the names and GO accession numbers of the ancestor biological process terms, as well as the child terms of primary metabolic process (GO:0044238) (a child term of "metabolic process") and immune system process (GO:0002376). The branches indicate ancestor-child relationships of primary metabolic process and immune system process terms. The bubble plots show the antidepressants for which the biological processes were identified as significantly enriched in at least one statin-antidepressant pair.

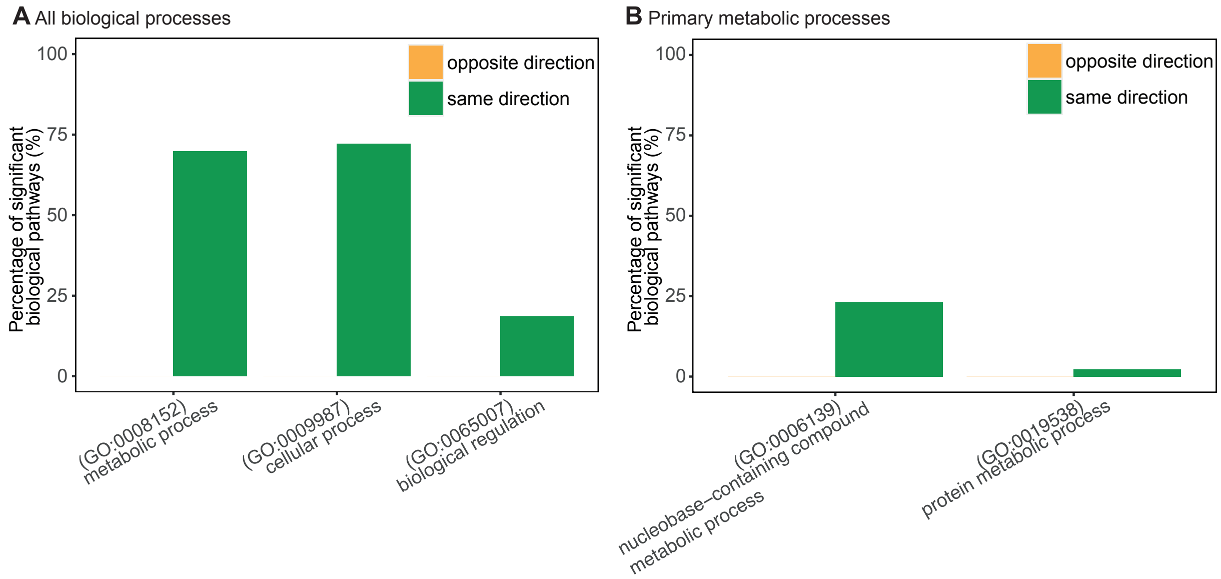

**Supplementary Figure 10.** Results of pathway enrichment analysis performed for genes perturbed by both statins and sirolimus. A total of 43 and 0 biological process terms were identified as significantly enriched amongst genes perturbed in the same and opposite directions, respectively, by statins and sirolimus. Enriched biological process terms were categorized into high-level (**A**) biological process and (**B**) primary metabolic process ancestor terms, and each term might be annotated with more than one ancestor. X-axis shows the names and GO accession numbers of the ancestor terms. Bar graphs show the percentages of significant GO biological process terms annotated with each ancestor term.

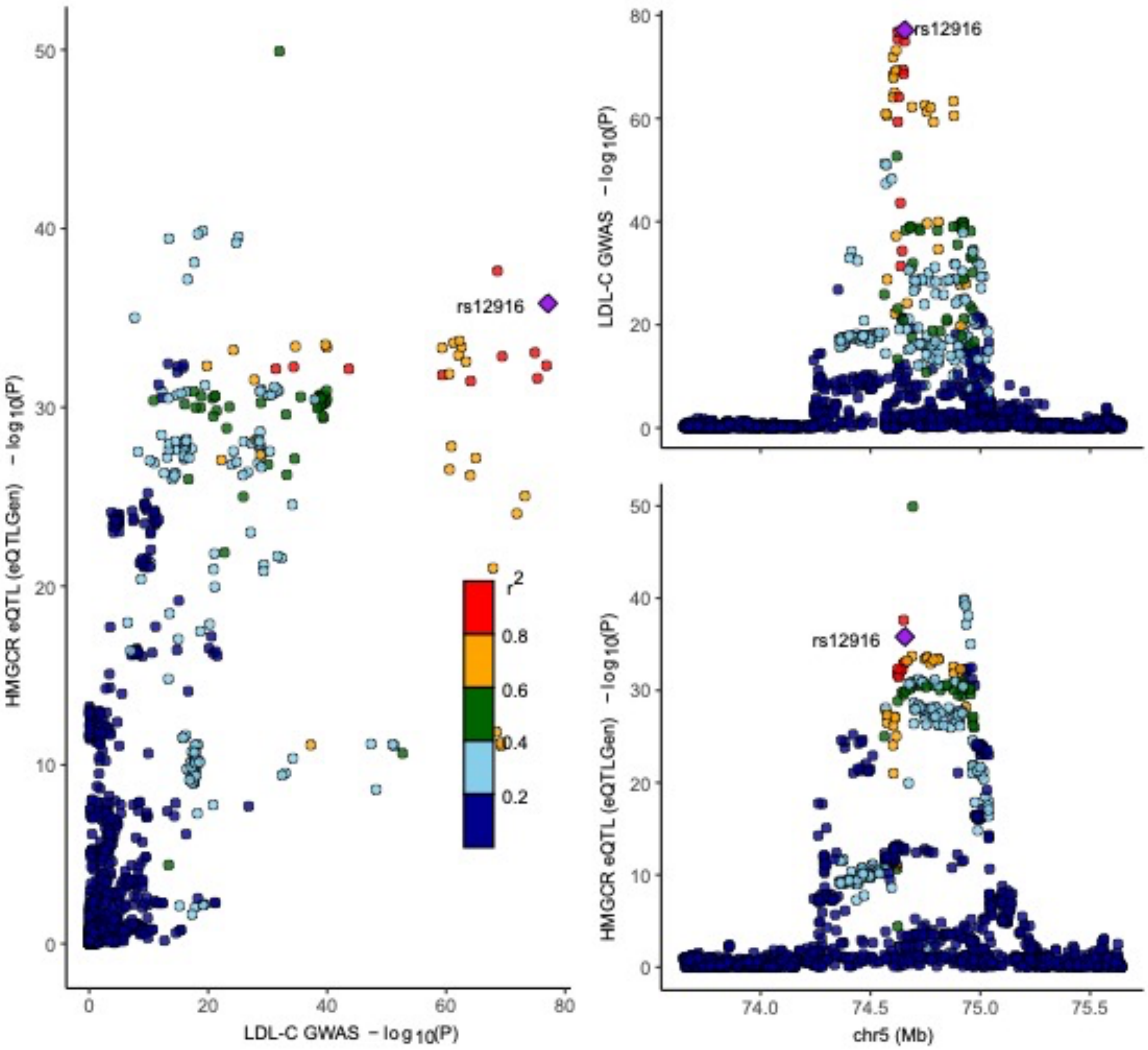

**Supplementary Figure 11.** LocusCompare plot of *HMGCR* eQTLs (eQTLGen) and LDL-C GWAS summary statistics. Each dot represents a SNP and is colored based on their degree of LD ($r^2$) with rs12916. The LocusZoom plots (right panels) show the significance (-$\log_{10}$P) of SNPs against their genomic locations. The scatter plot (left panel) shows the SNP significance in the eQTL dataset plotted against the SNP significance in the LDL-C GWAS dataset.

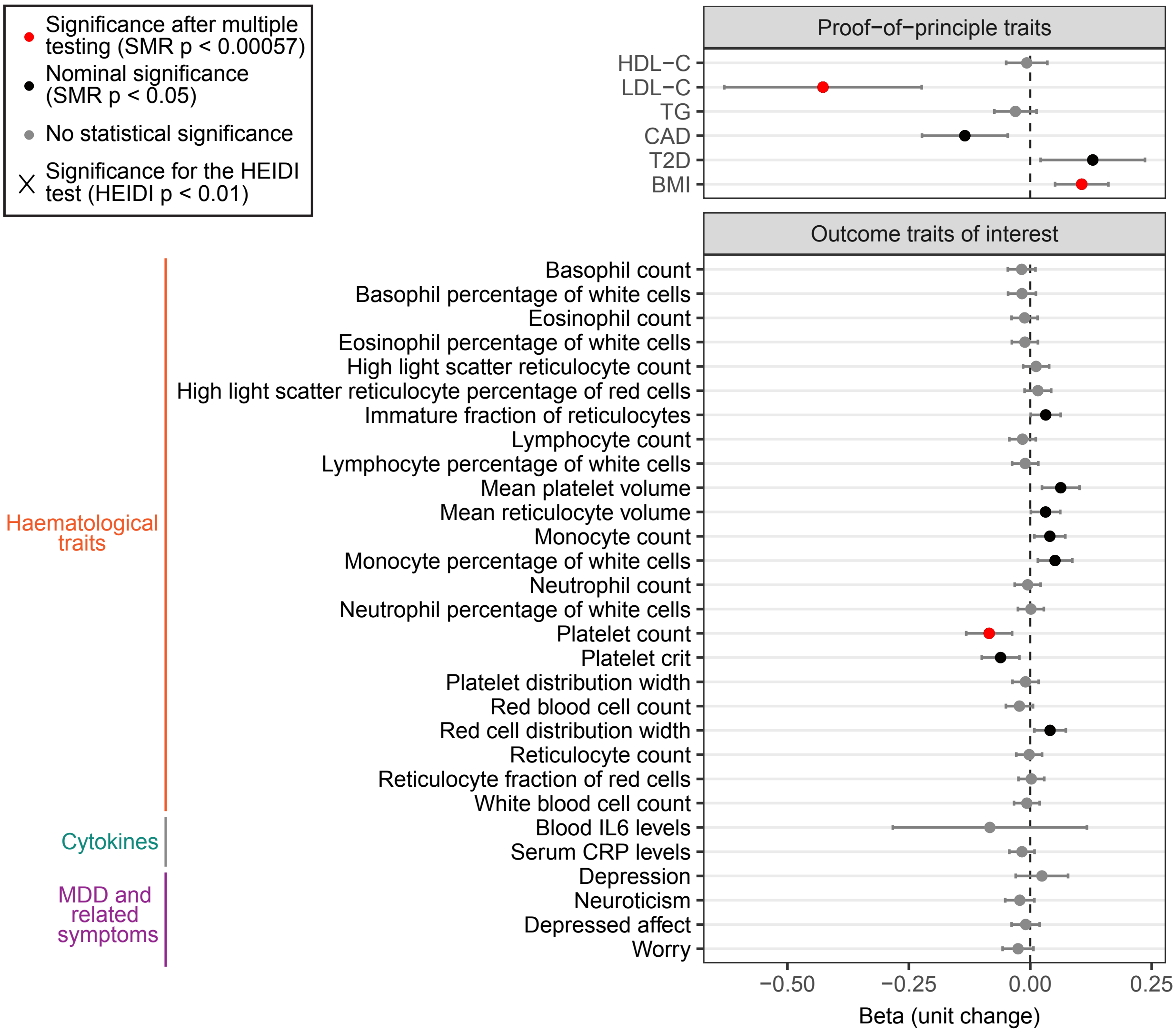

**Supplementary Figure 12.** MR analyses of *HMGCR* gene expression in the brain with various haematological, cytokine and depression-related traits. Dot plot shows the associations (beta) between gene expression and traits, and error bars show the 95% confidence intervals. The effect sizes are harmonized to represent the changes in trait per one standard deviation decrease in *HMGCR* expression. The beta values thus reflect unit changes in the outcome traits upon genetically proxied HMGCR inhibition. The units of beta values are not standardized. Red dots represent associations with statistical significance after multiple testing correction ($p < 0.00057$), and black dots represent associations with nominal statistical significance ($p < 0.05$). Associations with significant HEIDI p-values are marked by ×.
